# Affective Neuropsychiatric Symptom Metrics in the National Alzheimer’s Coordinating Center Dataset

**DOI:** 10.1101/2025.10.23.25338479

**Authors:** Daniel W. Fisher, Ronak Mehta, Christopher B. Morrow, Kathleen F. Kerr, Suman Jayadev, Kimiko Domoto-Reilly, Michael J. Schrift, Martin Darvas

## Abstract

**Background:** In dementia research, affective neuropsychiatric symptoms (NPS) – depression, anxiety, and apathy – remain understudied. Improving strategies to accurately identify clinically relevant NPS is essential for more robust research.

**Objectives:** We sought to determine how often objective metrics and clinical gestalt metrics agree on NPS presence or absence. We further sought to determine optimal cut-offs for affective NPS presence/absence using the Neuropsychiatric Inventory Questionnaire (NPI-Q) severity ratings.

**Methods:** We assessed agreement for NPS presence/absence among 5 different depression metrics, 4 anxiety metrics, and 2 apathy metrics via Jaccard indices using the National Alzheimer’s Coordinating Centers (NACC) dataset. Analysis included exploring different NPIQ severity rating thresholds of >0, >1, >2, and 0 and >1.

**Results:** NPIQ cut-off >1 for presence and =0 for absence of an NPS led to the best agreement with other metrics. However, there was poor agreement for NPS presence across depression metrics (6%) and across anxiety metrics (7%). Choice of metric could greatly skew the frequency of an NPS being present. All 3 affective NPS were more common in Lewy Body Disorder compared to Alzheimer’s Disease or Vascular Cognitive Impairment, regardless of metric.

**Conclusions:** Though NPIQ severity rating cut-off choice should depend on study design, using a severity score of >1 for presence and =0 for absence may best fit clinical gestalt for affective NPS. Lewy Body Disorders present with more affective NPS than other common dementia etiologies. Future consensus on criteria for depression and anxiety syndromes in dementia may improve their identification.

## Introduction

Neuropsychiatric symptoms (NPS) in dementia are one of the most important considerations for clinical management^1^, but receive much less attention than cognitive decline in molecular and cellular research^2^. This is despite the ubiquitousness of these NPS – occurring in 90% of patients Alzheimer’s Disease (AD) – and their direct link to decreased quality of life^3,4^, increased institutionalization, reduced independence^5,6^, and increased healthcare costs^7^. Further, NPS may be harbingers of neurodegenerative disease, as they present early in the disease course in a syndrome called Mild Behavioral Impairment (MBI)^8,9^. The advent of anti-amyloid therapies^10^ and specific biomarkers for dementia etiologies^11^ allow for disease modifying treatment and early detection, respectively; thus, identifying the earliest symptoms of neurodegeneration has never been more important. If NPS are identified before cognitive decline, one’s pretest probability may be elevated and facilitate biomarker confirmation, leading to earlier disease modifying therapy implementation and better outcomes^12^. “Affective” NPS, mainly depression, anxiety, and apathy, are present in over 50% of those with dementia^13^ and are the most common NPS in MBI^14^, making their recognition and accurate measurement of particular importance.

The difficulties with recognition and measurement of these NPS is one reason they are likely to be less well studied than cognitive decline, which benefits from robust and quantitative neuropsychological testing tools^15,16^. Though many clinical instruments have been developed to identify and measure NPS^16^, the NPI and the shortened NPIQ are arguably the most commonly used tools to assess NPS across cohort studies, mechanistic studies, and >350 clinical trials^17,18^. The original NPI recommended certain NPS may only be clinically relevant at certain severity/frequency cut-off scores (0-12), with >5 suggested for dysphoria/depression^19^. Many studies have erred on the side of caution, using a score >3 for the presence of clinically-relevant NPS^20–22^, including CATIE-AD, where NPI > 3 for agitation domains were necessary for study inclusion^23^. Despite this, the NPIQ measures only severity (0-3) and has no parallel recommendation for which scores indicate NPS presence or absence^24^. Most studies define a severity >0 as NPS presence^25–30^, but others use a threshold of >1^31–33^. This gap in how to interpret the NPIQ is especially salient, as many large cohort studies rely on this instrument for predictions about NPS prevalence throughout the disease course.

In an attempt to address this gap, we analyzed affective NPS measurements from one of the largest studies of aging and dementia from the National Alzheimer’s Coordinating Centers (NACC), which has prospectively followed participants from over 42 different sites. The harmonized data, known as the Uniform Data Set (UDS), contains a multitude of metrics to identify depression, anxiety, and apathy, including the NPIQ, Geriatric Depression Scale (GDS), and different clinical gestalt metrics. Given these paired objective (NPIQ, GDS) and clinical gestalt metrics, we sought to determine how these two types of metrics agreed across participants. We also investigated which NPIQ cutoffs had the best agreement with other metrics, allowing for an exploratory investigation into which NPIQ severity cutoffs match clinician’s evaluations of clinically relevant NPS. Finally, we determined how different metrics influenced the estimated frequency of these symptoms across cognitive stages (Cognitively Normal, CN; Mild Cognitive Impairment, MCI; and Dementia) and clinical etiologies (AD; Frontotemporal Dementia, FTD; Lewy Body Disorder, LBD; and Vascular Cognitive Impairment, VCI).

## Methods

### Clinical Case Series and Dataset

The NACC UDS is a dataset of clinical variables collected from 2005 to 2021 from 42 past and present ADRCs funded by the National Institute of Aging according to the individual recruitment strategies and priorities of each individual center. Though each ADRC reporting to NACC has unique missions and recruitment strategies/goals, all participants in their Clinical Core have a unified set of data collected in the UDS. While the focus of all centers is to study neurodegenerative dementias, and while the UDS collects the same clinical variables on every participant, there is significant heterogeneity in patient populations and clinical focus that may lead to selection bias. Though the participants are tracked longitudinally and remain in the study if meeting the criteria for each center, these differences necessitate this dataset to be considered a large case series as opposed to a true prospective cohort – thus terms like ‘prevalence’ are less applicable than ‘frequency of symptoms’. In addition, for the UDS versions 1-3 used to collect NPS data, standardization of criteria for each NPS and method of collecting each NPS may not be present for all data (discussed more below). Data were obtained on 9/6/2024 and consisted of 192,088 clinical visits across 51,836 unique participants. Because our aim was to study an aging population with a high rate of spontaneous neurodegenerative disease, we included subjects 60 years or older, leaving 177,960 clinical visits (92.6% of total) across 47,256 participants (91.2% of total). The UDS was structured to encompass 4 sections, A-D, each with additional subsections. All participants (and their study partners/co-participants) provided written informed consent under protocols approved by each ADRC’s Institutional Review Board. Data are collected annually (or as specified) and submitted to NACC under a Data Use Agreement.

### Cognitive Status and Etiology

Participants were categorized based on two different features: cognitive status and primary etiology. The cognitive status was diagnosed as Cognitively Normal (CN), amnestic or non-amnestic Mild Cognitive Impairment (MCI), Dementia, or cognitively impaired but does not meet criteria for MCI (impaired-not-MCI), with this latter group being excluded from analysis. Cognitive status was diagnosed in section D1 either by consensus panel or single clinician if no panel was available. In the UDS 3, CN was defined as “global CDR = 0 and/or neuropsychological testing within normal range” as well as “normal behavior”, deemed behavior that is “not sufficient to diagnose MCI or dementia due to FTD or LBD”. MCI was defined as notable impairment reported from the participant or co-participant, objective impairment in at least one cognitive domain, and preserved independence in functional abilities. Dementia was defined as having cognitive or behavior symptoms that interfere with the ability to function, is a decline from previous functional levels, are not explained by delirium or psychiatric disorder, include cognitive impairment by history and objective testing, and have clear impairment in one of the following domains: memory, executive function, visuospatial abilities, or language. Similarly, a change in personality, behavior, or comportment was considered another domain for impairment.

Primary etiologies were similarly diagnosed by consensus panel or single clinician and were based on clinical presentations matching syndromic presentation; biomarkers and imaging were not supposed to be used to consider the diagnosis – with the exception of VCI. Four primary etiologies were included in the analysis: AD, FTD, LBD, and VCI. AD was defined by 2011 NIA-AA criteria for AD dementia^34^. Behavioral variant FTD was defined by the International consensus criteria for behavioral variant FTD (FTDC) and FTD with Motor Neuron Disease was combined with this classification into an FTD category. Primary progressive aphasias, cortical basal degeneration, primary supranuclear palsies, and amyotrophic lateral sclerosis diagnoses were sparse in the database and considered too divergent from FTD to add to this category; subsequently, these etiologies were not analyzed. LBD was diagnosed by the Revised 2017 Criteria for the clinical diagnosis of Dementia with Lewy Body^35^. VCI was diagnosed by evidence of significant vascular brain injury confirmed by clinical evidence, such as presentation consistent with stroke, or neuroimaging studies. Further, to be considered the primary etiology, there needed to be one or more of the following: A temporal relationship between symptomatic stroke and cognitive decline, imaging evidence of cystic infarctions localized to a cognitive network, or cystic infarct and extensive white matter intensities with concomitant executive dysfunction.

### Neuropsychiatric Metrics

The UDS collects data on NPS in 5 places: Subject Health History (Form A5), named EMR/Report hereafter; the NPIQ (B5; Form B5), named NPIQ; Geriatric Depression Scale (GDS; Form B6); Clinician Judgment of Symptoms (Form B9), named Clinician-rated; and Clinician Diagnosis (Form D1), named Consensus-rated. For depression, all five metrics (EMR/Report, Clinician-rated, Consensus-rated, NPIQ, and GDS) were available. For anxiety, all sections except the GDS were available (EMR/Report, Clinician-rated, Consensus-rated, and NPIQ). For apathy, only Clinician-rated and NPIQ were available. Each metric has slightly different language and definitions for this dataset. For all metrics, if an answer was not filled out or marked unknown, the response was marked ‘NA’ and that entry was not used in the subsequent analysis.

For EMR/Report, data was gathered by the intake clinician using “ADRC scheduling records, subject interview, medical records, and proxy co-participant report.” For depression, the question could be scored as 1) active in the last two years, not active in the last two years, or unknown as well as 2) depression episodes more than two year ago, not present, or unknown. Further instructions in the UDS coding guidebook stated “During the interview, confirm with the subject and/or informant that the reported history of depression was based on a diagnosis and/or treatment by a physician/clinician.” For this analysis, depression presence on EMR/Report was defined as answering ‘yes’ to active in past two years only. Anxiety presence was indicated as ‘yes’, ‘no’, or ‘unknown.’

The NPIQ is a quick checklist of 12 NPS that is designed to be asked to an informant^17,24^, who rates each NPS as present/absent over the last month, and if present, as mild (1), moderate (2), or severe (3) severity. While the instructions describe only including “changes that have occurred since the patient first began to experience memory (i.e., cognitive) problems,” this is not always followed routinely by all clinicians (personal correspondence). Further instructions in the coding guidebook stated “Please report any behaviors or symptoms that were present within the last month, ignoring behaviors and symptoms that are usual for the subject and have been customary throughout his/her life.” For depression/dysphoria, the informant is asked “Does the patient seem sad or say that he/she is depressed?. For anxiety, the informant is asked “Does the patient become upset when separated from you? Does he/she have any other signs of nervousness such as shortness of breath, sighing, being unable to relax, or feeling excessively tense?”. For apathy/indifference, the informant is asked “Does the patient seem less interested in his/her usual activities or in the activities and plans of others?” While the presence and absence of an NPS can be defined using the presence/absence criteria, different cohort studies sometimes use different severity cut-offs^31–33^, possibly being more stringent to increase the likelihood of identifying clinically relevant symptoms. To explore an optimal cut-off in this dataset, the NPIQ was first converted to a score of 0 (absent) to 3, with 1-3 corresponding to the severity measures. Then, three potential cutoffs for the NPIQ were considered in the analysis, with a score above the threshold representing an NPS as present and below the threshold as absent: NPIQ > 0, NPIQ > 1, and NPIQ > 2. A fourth cutoff where an NPS was present with a NPIQ > 1 and absent with an NPIQ = 0 was also analyzed. With this 4^th^ cutoff, scores where NPIQ = 1 were deemed ambiguous and were ignored in the subsequent analyses. In each case, this led to a reduction in the number of analyzed participants, and the remainder of the total for each NPS is as follows: 85% for depression, 87% for anxiety, and 89% for apathy.

The GDS was designed to rate the likelihood of depression in older age populations and was created to avoid confounding effects of somatic symptoms that may occur in this population^36^. The GDS is filled out by the clinician based on the responses from the participant. While originally 30 questions with yes (1) and no (0) responses, the UDS uses the shortened 15 question version, that has also been validated as reliable and internally consistent^37^. The most commonly used and recommended cutoff for the GDS is >5, representing the likelihood of depression being present. Based on a meta-analysis^37^, a pooled specificity of 0.89 and sensitivity of 0.77 corresponds to this cutoff. With a reported prevalence of depression at 11.5% from this meta-analysis, a 2×2 table of a positive and negative GDS measures compared to the gold standard DSM-V diagnosis could be created. This 2×2 table was used in later analyses to compare Jaccard indices for agreement between the gold standard and the GDS in this meta-analysis.

The Clinician-rated NPS is based on the judgment of the intake clinician and scored as either yes, no, or unknown. The most recent UDS 3 describes the instructions for this section, with language reproduced in this methods section as appropriate. For each NPS, the clinician is instructed to “indicate whether the subject currently manifests meaningful change in behavior.” For “depressed mood”, the clinician answers “has the subject seemed depressed for more than two weeks at a time, e.g., shown loss of interest or pleasure in nearly all activities, sadness, hopelessness, loss of appetite, fatigue?” For anxiety, it asks “for example, does s/he show signs of nervousness (e.g., frequent sighing, anxious facial expressions, or hand-wringing) and/or excessive worrying?” For apathy/withdrawal, it asks “has the subject lost interest in or displayed a reduced ability to initiate usual activities and social interaction, such as conversing with family and/or friends?” A score of ‘yes’ on this section indicated the presence of the NPS and a ‘no’ indicated the absence of an NPS.

For the Consensus-rated metric, the presence or absence of an NPS was derived from one of three methods: From the clinician’s assessment, by “a formal consensus panel if the diagnosis was made by a group of clinicians (e.g., neurologists, neuropsychologists, geriatricians) who convene on a regular or semi-regular basis to discuss and decide upon the final diagnosis”, and by “an ad hoc consensus group (e.g., two or more clinicians or other informal group.” For this analysis, the group or clinician responsible for assessing this metric was not considered, instead, defining each NPS based on whether it was answered ‘yes’ as present or ‘no’ as absent. The NPS were named “Active Depression” and “Anxiety Disorder” in this section. Further instructions of how to define these symptoms were not provided for the individual clinician or consensus group in the UDS form; however, in the coding guidebook, guidance on making these diagnoses are given. For depression, it says “consult the Diagnostic and Statistical Manual of Mental Disorders regarding the diagnosis of depression,” and for anxiety it says “Consult the Diagnostic and Statistical Manual of Mental Disorders regarding the diagnosis of [Anxiety Disorder].” It is not clear for either diagnosis (neither “Active Depression” nor “Anxiety Disorder” is a DSM-V diagnosis) what formal DSM-V diagnosis should be used, such as Major Depressive Disorder, Pervasive Depressive Disorder, Adjustment Disorder with Depressed Mood, etc; Generalized Anxiety Disorder, Panic Disorder, Post-Traumatic Stress Disorder, etc.

Entries with codes for unknown or unavailable were converted to ‘NA’, and these entries were ignored for each analysis of these variables specifically. Not all NPS metrics were asked consistently from UDS 1-3. For depression in the EMR/Report, this variable was not obtained in follow-up visits in the UDS3. For anxiety in the EMR/Report, Clinician-rated, and Consensus-rated, this variable was not obtained in UDS 1 or 2. Due to these changes, data for EMR/Report depression was available in 63% of entries, EMR/Report anxiety in 9% of entries, Clinician-rated anxiety in 42% of entries, and Consensus-rated anxiety in 46% of entries. GDS was present for 86% of entries while all other metrics were available >93%.

### Statistics

To avoid confusion, we refer to the individual NPS determinations in the UDS as “metrics” (EMR/Report, Clinician-rated, Consensus-rated, NPIQ, GDS), whereas statistical summaries of agreement are noted as “measures”. Regarding these measures, we first note particular considerations in this analysis that differ from inter-rater reliability (IRR)-type analyses, which would otherwise motivate kappa statistics such as Cohen’s kappa ^38^. Our goal is to quantify the internal consistency between fixed, possibly dependent entities – the various NPS metrics in the UDS – which are seen as a property of the metrics themselves. By contrast, IRR statistics often measure the internal consistency of possibly varying, independent entities – such as survey respondents – which are seen as a property of the units being measured. Moreover, as there is not a “gold standard” across the various NPS metrics, common approaches to determining sensitivity, specificity, and Area-Under-the-Curve analyses are unfeasible. Instead, positive percent agreement (PPA) or negative percent agreement (NPA) are more applicable. Considering a 2 x 2 contingency table:

|  | Positive<br>(Metric 2) | Negative<br>(Metric 2) |
| --- | --- | --- |
| Positive (Metric 1) | $a$ | $b$ |
| Negative (Metric 1) | $c$ | $d$ |

, PPA is defined as *a*/(*a* + *b*) whereas the NPA is defined as *d*/(*c* + *d*). While ‘percent agreement’ has a clear interpretation, this metric can be misleading in situations with very high or very low prevalence because overall agreement will be high by chance Additionally, we wish to consider agreement across more than two metrics for a joint representation of agreement. Therefore, to account for the lack of gold standard and to generalize to >2 metrics, we employ the Jaccard similarity index^39^. Precisely, we define the symmetric measures

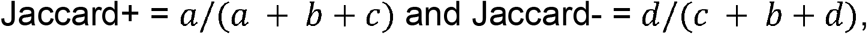

which measure agreement only on instances that are labeled as positive or negative by either metric. This agreement index has been employed both in traditional and modern statistical applications^39,40^. Note that the Jaccard index can be computed by

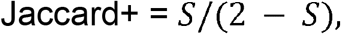

where *S* denotes the harmonic mean between the two PPA’s generated by treating either metric as the gold standard (i.e. the Dice-Sørensen coefficient). This further motivates the use of the measure as a symmetrized PPA, with an analogous relation holding for Jaccard- and NPA. To generalize the Jaccard+ to beyond two metrics, we take the ratio of the number of examples labeled positive by *all* metrics over the number of examples labeled positive by *any* metric; the Jaccard-is generalized in an analogous way. While uncertainty quantification for Jaccard statistics can be computed via bootstrap resampling^41^, the sample sizes for these calculations are extremely large (tens of thousands), so we consider the percentages as accurate estimates. For completeness, we also calculated the Cohen’s Kappa using the ‘psych’ package^42^.

A Youden’s J metric is often calculated to capture the performance of a diagnostic test and determine the optimal thresholds to maximize sensitivity and specificity. Usually calculated as J = Sensitivity + Specificity – 1, a similar Youden’s “J-like” statistic was calculated to evaluate optimal NPIQ threshold, calculated as J-like = Jaccard^+^ + Jaccard^-^ - 1. All statistical analyses were performed in R 4.3.2^43^.

## Results

### Descriptive statistics for dataset

To study a geriatric population and increase the likelihood of non-familial neurodegenerative disease, analyses were restricted to participants greater than 60 years old, resulting in 177,960 clinical visits (92.6% of total) across 47,256 participants (91.2% of total). Descriptive statistics of the clinical variables for the whole dataset and subsets by cognitive status were compiled (**Supplementary Table 1**). The average age at each study visit was 76.1 years of age (standard deviation 8.4); gender was 58.1% female; education level 15.6 years of school (standard deviation 3.3); *APOE e4* carrier rate was 38.0% with *APOE e2* carrier rate at 12.9%; 60.5% were married; and participants self-identified by the following NIH race definitions: 0.4% American Indian/Alaska Native, 2.5% Asian, 2.8% Biracial, 12.4% Black, 81.9% Caucasian, 0.1% Native Hawaiian Pacific Islander.

### NPIQ threshold agreement

First, the NPIQ metrics at different thresholds were analyzed for Jaccard+ and Jaccard-agreement with the other 4 metrics (**Figure 1A-C;** for Jaccard indices and PPA/NPA, see **Supplementary Figure 1**). As might be expected across all NPS metrics, the least stringent >0 threshold led to a higher Jaccard+ and lower Jaccard-than the most stringent >2 threshold. However, the approach of calling an NPS present with NPIQ > 1 and absent when =0 tended to yield improvements in agreement across both Jaccard+ and Jaccard-indices. Specifically, for depression, the NPIQ > 1 and =0 threshold had the highest Jaccard+ scores for agreement with GDS and EMR/Report metrics – even higher than > 0. For the clinician-rated and consensus-rated agreements, the > 1 and =0 threshold had the second highest Jaccard+ while the > 0 and > 1 had the highest scores, respectively. For depression and the Jaccard-score, the >1 and =0 threshold had the highest scores for the clinician-rated and consensus-rated metrics. For the GDS metric, the >1 and =0 threshold had the second highest rated score – with the >2 having the highest – but was only the third highest for the EMR/Report metric.

**Figure 1:**
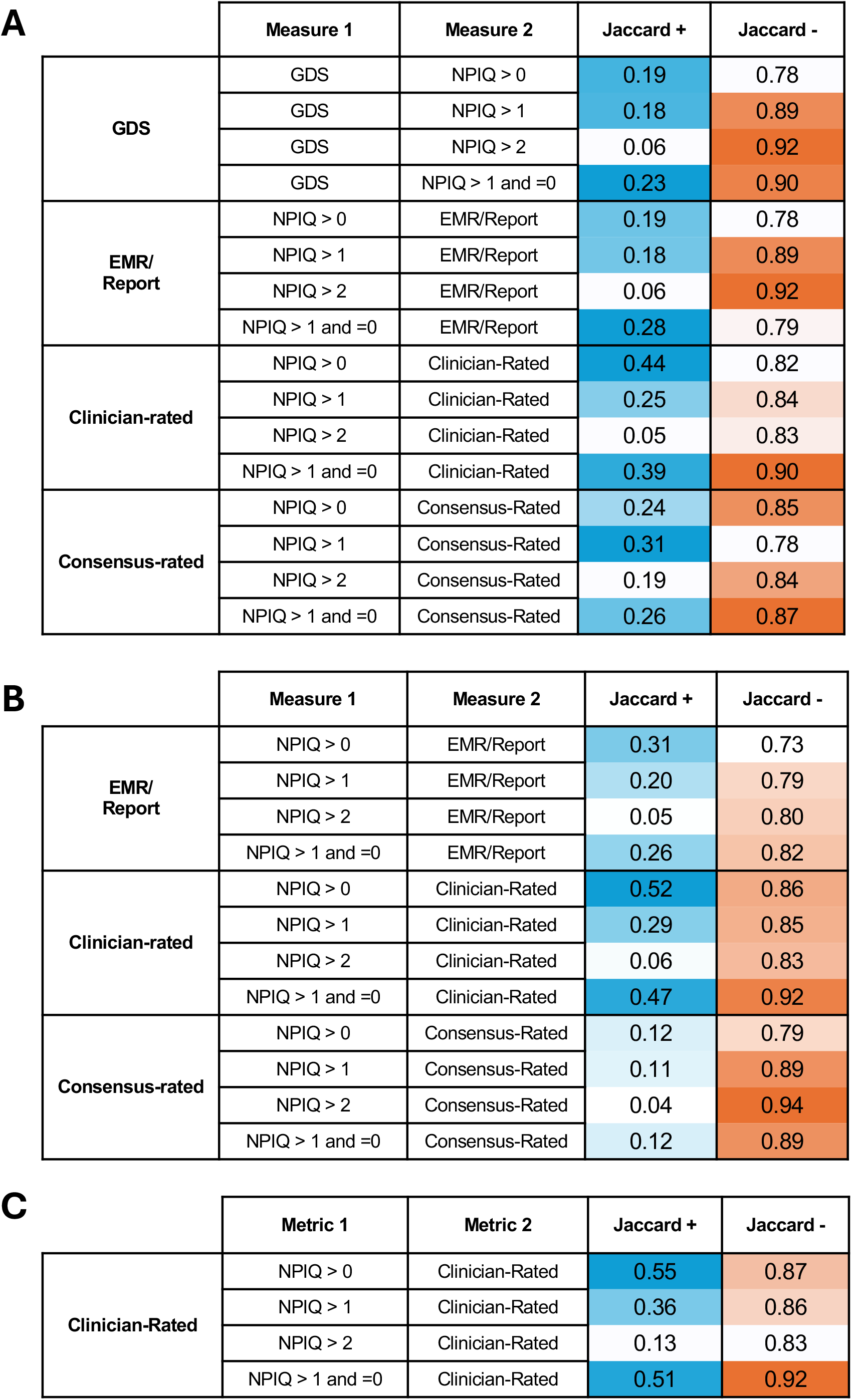
Determining an optimal NPIQ severity threshold for clinically relevant Affective Neuropsychiatric Symptoms. Jaccard indices detail agreement across two metrics for when a NPS is present (Jaccard+) and when it is absent (Jaccard-). Degree of agreement is depicted as a number of 0 (no agreement) to 1 (total agreement) with increased color intensity for agreement for presence (blue) and agreement for absence (orange). Results of agreement for different NPI thresholds are shown for **A)** Depression, **B)** Anxiety, and **C)** Apathy. Across all metrics and NPS, the NPIQ > 1 and =0 threshold tended to lead to the best maximization of Jaccard+ and Jaccard-scores and was therefore used as the subsequent threshold for the NPIQ across the rest of the agreement analyses, unless otherwise specified. Neuropsychiatric Symptom, NPS; Geriatric Depression Scale, GDS; Neuropsychiatric Inventory Questionnaire, NPIQ; Electronic Medical Record; EMR.

For anxiety, the >1 and =0 threshold had the 2^nd^ highest Jaccard+ score for all metric pairs with the >0 threshold having the highest. For the clinician-rated and EMR/Report metrics, the >1 and =0 threshold had the highest Jaccard- and was second for the Consensus-rated metric behind the >2 threshold.

For apathy, the >1 and =0 threshold had the second highest Jaccard+ score behind the >0 threshold, and the highest Jaccard-score.

Though imperfect, to get a sense of an aggregated statistic for Jaccard+ and Jaccard-score, a Youden’s “J-like” statistic was calculated using Jaccard+ and Jaccard-scores instead of using sensitivity and specificity (**Supplementary Figure 2**). In all cases, the >1 and =0 threshold led to a higher Youden’s J-like score across NPS. Cohen’s Kappa was also calculated (**Supplemental Figure 3**), though these values can be influenced by the prevalence of positive and negative ratings, as Kappa adjusts for chance agreement which itself depends on marginal distributions. In nearly every case, the Kappa values for the NPIQ thresholds of >1 or =0 and >0 were similar with overlapping confidence intervals, whereas alternative thresholds generally yielded lower Kappa values. Exceptions included comparisons of GDS with NPIQ in Depression and Consensus-Rated with NPIQ in Anxiety, where Kappa was higher for the >1 or =0 threshold. Conversely, for the EMR/Report and NPIQ comparison in Depression, the >0 threshold yielded a slightly higher Kappa.

**Figure 2:**
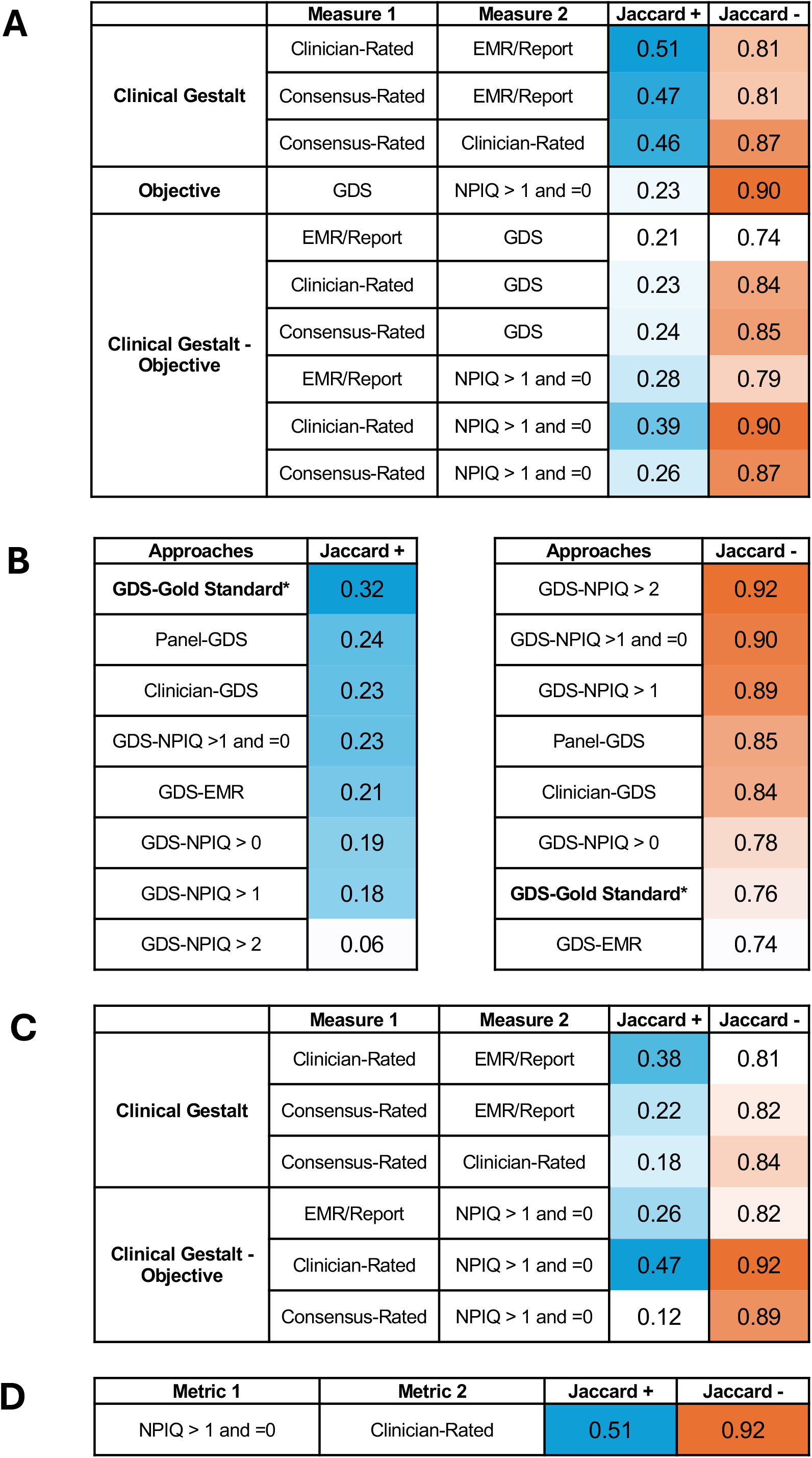
Agreement across NPS metrics. Jaccard indices detail agreement across two metrics for when a NPS is present (Jaccard+) and when it is absent (Jaccard-). Degree of agreement is depicted as a number of 0 (no agreement) to 1 (total agreement) with increased color intensity for agreement for presence (blue) and agreement for absence (orange). **A)** Agreement amongst the 5 depression metrics ordered by clinical gestalt pairs (EMR/Report, Clinician-rated, Consensus-rated), objective clinical tool pairs (NPIQ, GDS), and clinical gestalt-objective pairs. **B)** Gold-standard clinician diagnosis of Major Depressive Disorder was compared to GDS performance (bolded and starred) and compared to depression metrics in the UDS. **C)** Agreement amongst the 4 anxiety metrics, **D)** Agreement amongst the 2 apathy metrics. Neuropsychiatric Symptom, NPS; Geriatric Depression Scale, GDS; Neuropsychiatric Inventory Questionnaire, NPIQ; Electronic Medical Record; EMR.

The joint agreement across all metrics was also assessed, which is an extension of the Jaccard indices but with inclusion of 2 or more metrics (**Supplementary Figures 4, 5**). Similar to the pairwise Jaccard indices, the >1 and =0 threshold was consistently ranked 2^nd^ highest across depression and anxiety on both the positive and negative joint indices, sometimes showing higher agreement with more than one clinical gestalt metric (EMR/Report, Clinician-rated, Consensus-rated) than a pairwise comparison using a less optimal threshold.

Though altogether the >1 and =0 threshold maximized the likelihood of Jaccard+ and Jaccard-agreement scores across NPS and metric pairs, a cost came at the number of clinical visits that could be included in the analysis. Specifically, for those with a NPIQ of 1, the presence or absence of an NPS was termed ‘ambiguous’ or ‘indeterminate’, thus reducing sample size if you exclude this intermediate category. This led to 15.3% of entries being ambiguous for depression, 13.1% being ambiguous for anxiety, and 10.5% being ambiguous for apathy.

### Agreement across all metrics

For the 5 metrics available for depression (**Figure 2A**), Jaccard+ scores ranged from 0.05 to 0.51 while Jaccard-ranged from 0.74 to 0.92. When the rate of a particular positive metric is relatively low, such as with the NPS in this case-series^44^, Jaccard+ scores will be much lower on average than Jaccard-, as each disagreement will have a larger impact on the Jaccard+ due to fewer overall times the metric comes up positive. Focusing on the Jaccard+ indices for depression, greater agreement was noticed for the three metrics that relied on clinical gestalt: EMR/Report, Clinician-Rated, and Consensus Rated (range 0.46 – 0.51). By contrast, the metrics that relied on objective cut-offs, the NPIQ and GDS, had much poorer agreement based on Jaccard+ scores (range 0.06 – 0.23). When one of the objective metrics was paired with one based on clinical gestalt, Jaccard+ agreement still tended to be poorer (range 0.05 – 0.44).

Similarly, when comparing the Jaccard-scores, the metrics based on clinical gestalt showed poorer agreement (range 0.81 – 0.87) than the objective metrics (range 0.78 – 0.92), though the NPIQ > 0 cutoff with a Jaccard-of 0.78 greatly skewed the range for the objective measures towards lower values. For the Jaccard-agreement between objective metrics and clinical gestalt, the level of agreement was variable (range 0.74 – 0.92), though Jaccard-agreement tended to be higher for the NPIQ and clinical gestalt pairs (range 0.78 – 0.92) than the GDS and clinical gestalt pairs (range 0.74 – 0.85).

When looking at joint comparisons over two or more metrics (**Supplementary Figure 4B, D**), the three clinical gestalt metrics had higher positive agreement than almost all the combinations of the objective metrics. For negative joint agreement, some of the combinations of three metrics including objective and clinical gestalt had higher agreement than some of the pairs of clinical gestalt metrics, especially when including EMR/Report. Overall, for depression, this suggests that clinical gestalt may be more likely to overcall an NPS being present or that the objective metrics are not as sensitive to clinically relevant but lower severity cases.

Because the GDS has been studied in the context of a gold standard – clinician-made diagnosis of Major Depressive Disorder via the DSM – the levels of agreement between the GDS and all other metrics can be put in context of the performance of the GDS with this gold standard (**Figure 2B**). Using a meta-analysis of GDS performance^37^, Jaccard+ and Jaccard-indices could be calculated from the sensitivity, specificity, and prevalence across multiple studies. For the Jaccard+ score, the GDS paired with the gold-standard was a 1/3 higher than the next best Jaccard+, which was for GDS and the Consensus-rated pair. As the prevalence of 10.5% in the meta-analysis was less than the projected prevalence using the average of the Consensus-rated and GDS metrics in this dataset (11.5%), a difference in prevalence alone could not explain the gap between these two Jaccard+ indices. By contrast, the Jaccard-score for the GDS and gold standard pair performed about 15% lower than the highest Jaccard-score, which was for the NPIQ and GDS. Again, the prevalence of MDD by the gold standard was higher than the rate of NPS being absent by the average of the NPIQ and GDS metrics (8.5%). These data suggest that when used in the context of this case-series, the GDS may not perform with the same accuracy as in other studies in the literature or that the construct of depression as an NPS in dementia differs from the DSM-based Major Depressive Disorder diagnosis.

Results were similar when a Cohen’s Kappa was calculated for the GDS, where the next highest Kappa – again, >1 and =0 threshold – was about 20% lower than compared to the gold standard (**Supplementary Figure 3F**). As this metric scales with prevalence, it corroborates the findings for the Jaccard Indices.

For anxiety (**Figure 2C**), clinical gestalt tended to have lower Jaccard+ scores (range 0.18 – 0.38) than for depression and was less consistent in terms of being higher than the clinical gestalt-objective pairings (range 0.12 – 0.47). For Jaccard-scores, clinical gestalt was often lower (range 0.81 – 0.84) than the objective and clinical gestalt pairs (0.82 – 0.92). The highest Jaccard+ and Jaccard-scores were interestingly for the Clinician-rated and NPIQ metrics (Jaccard+ 0.47; Jaccard-0.92). Combinations with the Consensus-rated metric tended to have low Jaccard+ scores (range 0.12 – 0.22) with higher Jaccard-scores (range 0.82 – 0.89), but not always. This was partially driven by the lower frequency of present NPS by this metric, which was nearly half the predicted frequency of the next lowest metric (NPIQ >1 and =0) and a fourth of the highest metric (EMR/Report). Similarly for the joint comparisons (**Supplementary Figure 5B, D**), the Clinician-rated, NPIQ, and EMR/Report combination had a higher Jaccard+ than the Consensus-rated and NPIQ combination, while the combination of the Consensus-rated, Clinician-rated, and NPIQ had a higher Jaccard-compared to almost all the other pairwise combinations. Overall, while the Jaccard+ agreements were lower for anxiety, it appears that the Consensus-rated metric is particularly out of sync with the other metrics.

Apathy only had the Consensus-rated metric and NPIQ to compare, which had a Jaccard+ of 0.51 and a Jaccard-of 0.92 (**Figure 2D**). Both these scores were higher than the highest Jaccard+ scores for depression (0.39) and anxiety (0.47) and in line with the Jaccard-scores for depression (0.90) and anxiety (0.92).

Finally, the relative agreement of all metrics for a particular NPS was analyzed for depression (**Supplementary Figure 4B**,**D**) and anxiety (**Supplementary Figure 5B**,**D**). For the 5 depression metrics, the joint positive agreement was 0.06; that is, that all 5 metrics agree with each other only 6% of the time. Even excluding EMR/Report, which could consider depression over a longer time period than the other metrics, the joint agreement was only 0.08. In contrast, the negative joint agreement was still quite high for all 5 metrics at 0.72. For anxiety’s 4 metrics, the positive joint agreement was 0.07 while the negative joint agreement was 0.75. Again, while the overall low frequency of NPS across the dataset should be taken into account, recognition of anxiety and depression in this dataset is fully agreed upon less than 10% of the time, while only about 25% of the time is there a disagreement about whether an NPS is absent.

### Agreement across all metrics by cognitive status

It is a well-documented phenomenon that affective NPS increase as cognitive stage worsens from CN to MCI to dementia in most neurodegenerative disorders^45,46^, even if the degree of this increase is less clear. Concurrently, it would be expected for the Jaccard+ score to increase across dementia stages as the Jaccard-score would drop based on the increasing NPS frequency alone. For depression (**Figure 3A)**, the clinical gestalt metrics’ agreements jumped precipitously from CN to MCI (differences between cognitive statuses 0.12 to 0.48 across range of metric pairs) but not as much from MCI to dementia (difference -0.04 to 0.07). By contrast, there was a more modest increase amongst the objective metrics compared to the clinical gestalt pairs from CN to MCI (difference 0.08) and MCI to dementia (difference 0.01), with the combinations of objective metrics and clinical gestalt falling in between for CN to MCI (difference 0.07 to 0.16) and MCI to dementia (difference -0.02 to 0.09). It should be noted that clinical gestalt paired with the NPIQ tended to have better agreement than when clinical gestalt was paired with the GDS, as might be expected based on the intended target populations for both screening tests. For the Jaccard-indices, an opposite trend was noted. Specifically, amongst the clinical gestalt metrics, there was a modest reduction in the Jaccard-from CN to MCI (difference -0.08 to -0.02) and a similar modest reduction when going from MCI to dementia (difference -0.03 to -0.09). For the objective metrics, there were modest reduction from CN to MCI (difference -0.08) and MCI to dementia (difference -0.07). Interestingly, the Jaccard-tended to drop slightly more in the combinations of clinical gestalt and objective metrics from CN to MCI (difference -0.08 to -0.16) and MCI to dementia (-0.04 to -0.11). When taken in total, it seems that the Jaccard+ tended to have a greater rise when going from CN to MCI than the Jaccard-dropped across the same cognitive statuses, though the changes in both indices comparing MCI to dementia were similar. In addition, the Jaccard+ agreement for clinical gestalt tended to be higher than the objective measures or clinical gestalt paired with objective measures across cognitive stages while the same sized differences were less noticeable for the Jaccard-scores.

**Figure 3:**
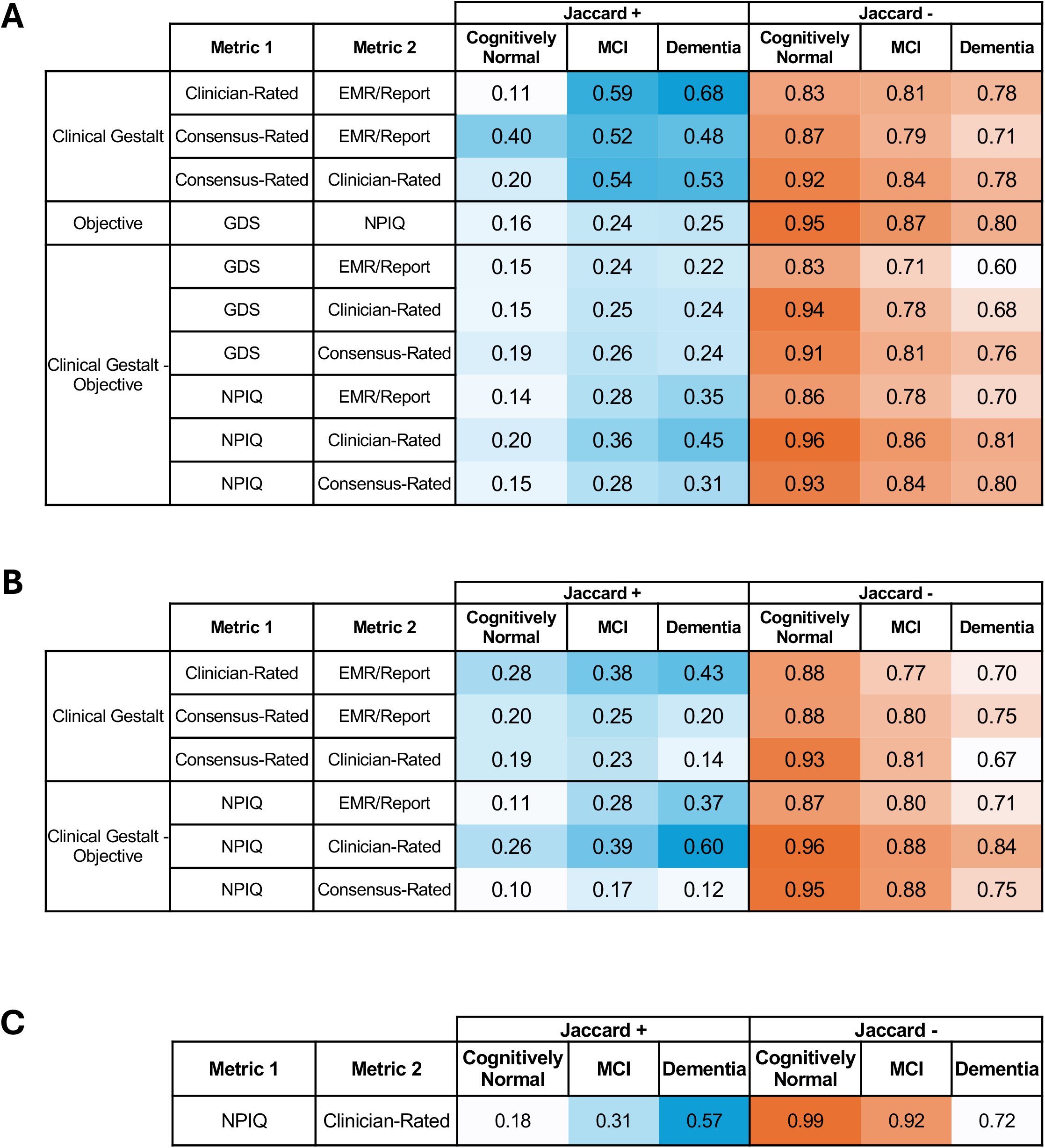
NPS agreement measures across cognitive statuses. Jaccard indices detail agreement across two metrics for when a NPS is present (Jaccard+) and when it is absent (Jaccard-), where the ratio depicts the total agreement over the total amount either metric suggests presence or absence. Jaccard indices were calculated from separate groups of participants who were diagnosed as cognitively normal, MCI, or dementia by the consensus panel. Degree of agreement for each metric pair is depicted as a number of 0 (no agreement) to 1 (total agreement) with increased color intensity for agreement for presence (blue) and agreement for absence (orange). Results of agreement for different NPI thresholds are shown for **A)** Depression, **B)** Anxiety, and **C)** Apathy. Neuropsychiatric Symptom, NPS; Geriatric Depression Scale, GDS; Neuropsychiatric Inventory Questionnaire, NPIQ; Electronic Medical Record; EMR; Mild Cognitive Impairment, MCI.

For anxiety (**Figure 3B**), there was also a pattern of increasing Jaccard+ agreement across cognitive statuses. Specifically, for clinical gestalt, there was a modest increase in Jaccard+ scores from CN to MCI (difference 0.04 – 0.11) and a less consistent change from MCI to dementia (difference -0.09 to 0.05). The objective measure combined with clinical gestalt tended to have better Jaccard+ score changes from CN to MCI (difference .07 to 0.17) and MCI to dementia (difference -0.05 to 0.21). While this was different than depression, but when comparing NPIQ agreements between NPS, anxiety and depression were similar across cognitive statuses – suggesting the lower objective agreements for depression were due to pairs with the GDS. For Jaccard-agreement, modest declines in scores were see from CN to MCI (difference -0.12 to - 0.07) and MCI to CN (difference -0.05 to -0.14) for the clinical gestalt measures, and similar modest declines were seen for the combinations of NPIQ and clinical gestalt from CN to MCI (difference -0.07 to -0.08) and MCI to CN (-0.04 to -0.13). When comparing the individual metrics, the Jaccard+ scores rose the least across cognitive statuses for the Consensus-related metric combinations while falling the most for the Jaccard-scores across cognitive statuses, suggesting that the Consensus-related metric may perform particularly poorly for identifying anxiety in more cognitively impaired individuals. Overall, the differences in the Jaccard+ and Jaccard-agreements were similar to depression, though the relative differences in objective measures and clinical gestalt combinations were not the same.

The differences in Jaccard+ and Jaccard-scores for apathy (**Supplementary Figure 4C**) mirrored those for the other NPS. Specifically, the Jaccard+ for apathy increased a moderate amount from CN to MCI (difference 0.13) and a large amount from MCI to dementia (difference 0.26). The Jaccard-for apathy similarly decreased a moderate amount from CN to MCI (difference -0.07) and a large amount from MCI to dementia (difference -0.20). When taken in context of all the affective NPS, this analysis suggests that while rising NPS frequency is likely to account for much of the differences in Jaccard+ and Jaccard-agreements, there may be subtle nuances in agreement that are dependent both on the specific NPS and cognitive status.

In almost every case, the Cohen’s Kappa for the Depression, Anxiety, and Apathy metrics followed the pattern of the Jaccard+ scores (**Supplementary Figure 6**). A similar pattern of more agreement in MCI and dementia than in CN was seen for Cohen’s Kappa across NPS, again suggesting that prevalence alone did not explain the increase in agreements suggested by the Jaccard+ measure. However, it remained rare that Cohen’s Kappa was higher than >0.6, with these notable instances being EMR/Report paired with Clinician-rated depression in MCI and Dementia (K_MCI_ = 0.64; K_DEM_ = 0.69), Consensus-rated and Clinician-rated depression in MCI (K_MCI_ = 0.62), and NPIQ and Clinician-rated anxiety in dementia (K_DEM_ = 0.66).

### Estimating NPS frequency by different metrics

Because the NACC dataset is better conceptualized as a very large case series than a true prospective cohort – due to different recruitment strategies across individual centers – determining NPS prevalence would be inappropriate. Still, understanding how NPS frequency estimates change by different metrics and cognitive status is helpful in understanding this population, as the large amount of biospecimens collected from this population are an impressively robust tool for future investigation of NPS. To explore this, we determined the frequency for when an NPS was present at each visit.

As might be expected by the poor Jaccard+ agreement, NPS frequency estimates showed wide ranges (**Figure 4A-C**). If including all NPIQ thresholds are included, the range of depression frequencies in the whole case-series was 0.01 to 0.23. When only including the NPIQ threshold >1 and =0, the range narrowed to 0.08 to 0.17. Anxiety frequency estimates ranged from 0.02 to 0.22 and narrowed to 0.05 and 0.20 when considering only the NPIQ >1 and =0 threshold. Apathy frequencies estimates ranged from 0.03 to 0.20, narrowing to 0.11 to 0.18 when only considering NPIQ >1 and =0. While the apathy estimates across the population were similar, even when restricting the analysis to NPQ >1 and =0, the top NPS frequency estimates with the highest metric were twice and four-times as much as the lowest metric for depression and anxiety, respectively.

As seen with the agreement analyses, metrics may have better or worse accuracies by cognitive status. Therefore, NPS frequency estimates for each metric were calculated in participants who were CN, had MCI, and had dementia; only the NPIQ >1 and =0 threshold was analyzed (**Figure 4D-F**). Depression frequencies differed over a 6-fold range for CN (range 0.03 to 0.18), a 2.9-fold range for MCI (range 0.11 – 0.32), and a 3-fold range for dementia (range 0.14 – 0.42). Objective metrics tended to have lower depression frequency estimates than clinical gestalt measures. Anxiety frequencies differed over a 4.7-fold range for CN (range 0.03 – 0.14), a 3.1-fold range for MCI (range 0.07 – 0.22), and a 5.8-fold range for dementia (range 0.06 – 0.35). The Consensus-related metric consistently estimated much lower anxiety frequencies than the other metrics. Apathy frequencies were the same for CN at 0.01, 2.3-fold different for MCI (range 0.06 – 0.14), and 1.4-fold different for dementia (range 0.34 – 0.48).

**Figure 4:**
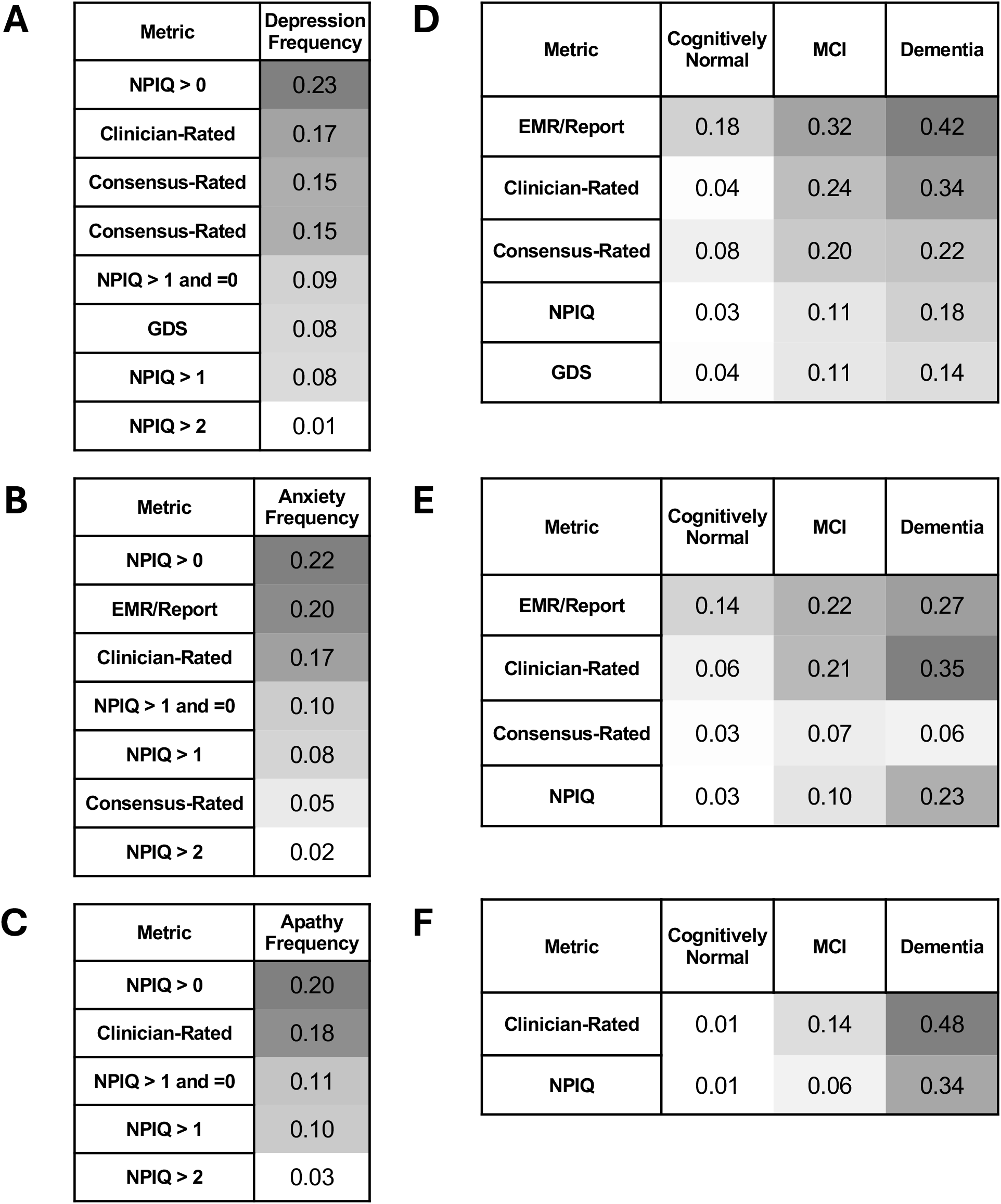
NPS Frequency by metric and cognitive status. The frequency of each NPS per clinical visit was estimated for **A)** Depression, **B)** Anxiety, and **C)** Apathy across NPIQ thresholds and the other metrics. Frequencies of each NPS were analyzed by cognitive status, with NPIQ threshold set at >1 and =0, for **D)** Depression, **E)** Anxiety, and **F)** Apathy. Neuropsychiatric Symptom, NPS; Geriatric Depression Scale, GDS; Neuropsychiatric Inventory Questionnaire, NPIQ; Electronic Medical Record; EMR.

Understanding NPS frequencies by different clinical syndromes can be helpful, as there are some contrasting data about the efficacy of certain pharmacotherapies for affective symptoms by underlying neurodegenerative etiology, especially for depression^47,48^. Therefore, NPS frequencies were calculated by clinical syndrome, specifically AD, LBD, FTD, and VCI in participants who had MCI or dementia (**Figure 5A-C**). Similar to the previous analyses, NPS frequencies varied greatly by metric, but some patterns emerged by etiology. There were similar rates of depression for AD (range 0.10 – 0.37), FTD (range 0.16 – 0.40), and VCI (range 0.13 – 0.38) but generally higher rates of depression for LBD (range 0.25 – 0.58). Anxiety similarly showed higher rates for those with LBD (range 0.07 – 0.41) than AD (range 0.06 – 0.29), FTD (range 0.04 – 0.31), and VCI (range 0.07 – 0.22). Apathy was unsurprisingly highest in FTD (range 0.51 – 0.60), where it is a core criteria, though rates were also higher in LBD (range 0.34 – 0.35) than in AD (0.22 – 0.35) and VCI (0.14 – 0.24). Still, whether across the whole case series, different cognitive statuses, or different etiologies, there were wide ranges in NPS frequencies when estimated by the different metrics included in this dataset.

**Figure 5:**
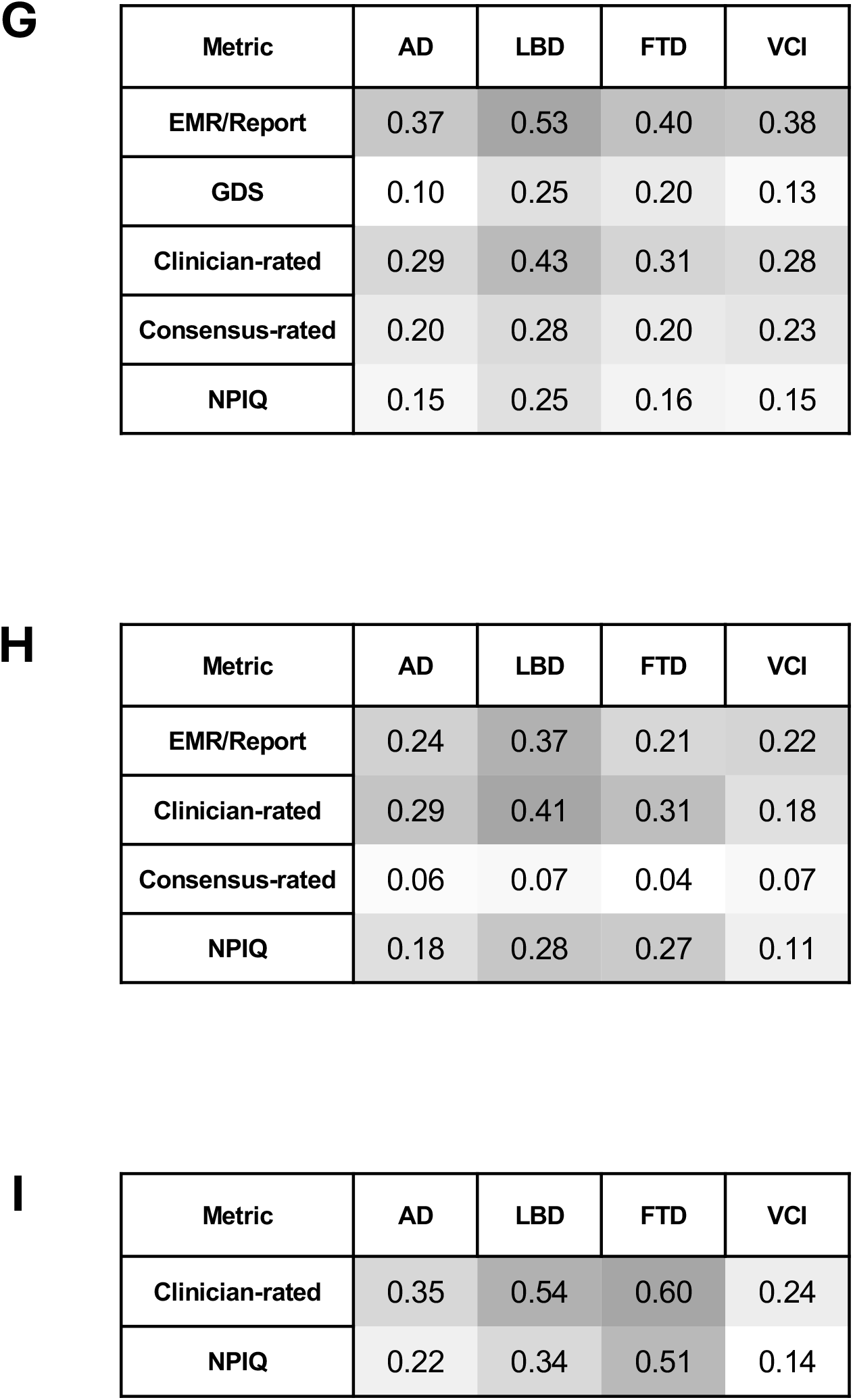
NPS Frequency by metric and etiology. Frequencies of each NPS were analyzed in MCI and dementia participants by primary etiology for **G)** Depression, **H)** Anxiety, and **I)** Apathy. Neuropsychiatric Symptom, NPS; Geriatric Depression Scale, GDS; Neuropsychiatric Inventory Questionnaire, NPIQ; Electronic Medical Record, EMR; Alzheimer’s Disease, AD; Lewy Body Disorder, LBD; Frontotemporal Dementia, FTD; Vascular Cognitive Impairment, VCI.

## Discussion

In this analysis of a large dataset of older adults from NACC, an NPIQ severity threshold of >1 for affective NPS presence and =-for NPS absence led to the best overall agreement with other NPS metrics for anxiety, apathy, and especially depression. However, there was generally poor agreement across the mix of clinical gestalt and objective metrics in this dataset, leading to a wide range of NPS frequency estimates depending on the metric employed. In general, metrics based on clinical gestalt had higher levels of agreement than those based on clinical gestalt-objective metric pairs. Despite these varied estimates, an increase in both NPS prevalence and metric agreement was seen as cognitive status worsened, and LBD tended to have a higher rate of all anxiety and depression than AD, FTD, or VCI.

### Choosing an NPIQ Threshold

With the growing popularity of the NPIQ over the fuller NPI, which involves a longer interview and more questions^24^, an increased need for clarity in the interpretation of the severity score for this instrument is essential – especially as it pertains to what NPS severity might best associate with the presence of clinical relevant affective NPS. Though no gold standard exists for establishing the presence of clinically relevant depression or anxiety in dementia – attempts have been made^49,50^ – and while the UDS 1-3 does not use the recently suggested apathy criteria for dementia^51^, the NACC dataset has at least one clinical gestalt diagnosis for each affective NPS. Throughout this analysis of agreement across these metrics, the best balance of agreements for when an NPS was present or absent was with the threshold NPIQ > 1 and =0, where NPIQ = 1 was treated as indeterminate. For depression, this seemed particularly salient, as there were even instances where agreement of depression presence was improved with the >1 and =0 threshold compared to the >0 or >1 threshold – such as agreement with GDS and EMR/Report – while the agreement for depression absence was always better with the >1 and =0 threshold. With mild dysphoria being common and possibly more related to adjustment disorder, which has different epidemiological and therapeutic implications than depression^52^, this more nuanced approach than simply defining depression as severity >0 may be especially important for depression compared to the other affective NPS. However, the ‘cost’ of this more restrictive threshold is in exclusion of those where NPIQ = 1. Still the risk of misclassification may increase noise beyond the gains in statistical power with inclusion of these participants. For any study, determining the tolerance for false positive and negatives is dependent on the study’s goals and resources, and the Jaccard+ and Jaccard-scores in this analysis may help guide researchers when choosing severity threshold for the NPIQ.

### Poor agreement across metrics

For depression and anxiety, the overall agreement amongst metrics was surprisingly poor: depression presence was agreed upon 6% of the time, and similar agreement for anxiety was 7%. Accordingly, the estimates of NPS frequency at each study visit were quite variable, at most being 6-fold different for anxiety for CN participants. Even apathy, which had two metrics, had a 2.4-fold difference in NPS frequency estimates for MCI.

One might hypothesize that differences in NPS stability over time – taking into account the slightly different time windows for each metric – or differences in clinician-, informant-, and self-ratings might drive this lack of agreement^27^. However, while it is true that NPIQ test-retest reliability was initially established over hours to weeks^19,24^, follow-up studies have noted these NPIQ assessments are stable over years-long periods^53^. In terms of the source of clinical information, the analysis comparing the GDS to clinician-rated, gold standard diagnoses suggests poor agreement of NPS presence still exists for this dataset compared to other studies – regardless of whether Jaccard indices or Cohen’s Kappa were employed. The reason for this poorer agreement is unclear, but calls attention to the lack of unified, gold standards for the diagnosis of affective NPS in dementia ^16,47,49–51^.

Focusing on depression as a prototypical affective NPS, there are multiple reasons why differentiating a specific depression syndrome in dementia might be important, as has been done for depression in other specific disease contexts, such as post-partum depression^47^. For one, clinical trial evidence demonstrates that serotonergic modulators do not separate from placebo for depression in dementia despite doing so in otherwise healthy older adults^48^. This may suggest either lack of viable, compensatory biochemical pathways for depression in dementia or completely different molecular mechanisms of depression pathogenesis with neurodegeneration. Further, the challenge of distinguishing apathy from anhedonia may be unique to neurodegenerative illnesses, leading to misdiagnosis of apathy as depression^47^. Indeed, some groups have suggested that focusing on dysphoria with the exclusion of anhedonia/amotivation may be a more accurate approach^47,54^.

Agreement for NPS presence generally improved with worsening cognitive status, and while the agreement for NPS absence worsened in parallel, the degree of worsening was of smaller magnitude. Increased NPS prevalence likely underlies a portion of this finding ^46^, but Cohen’s Kappa’s showed a similar trend. One possibility is that NACC clinicians may be more skilled at recognizing NPS in those who are cognitively impaired versus CN. Further, the NPIQ is designed for those with cognitive impairments, though it has been used in other settings^17^. Interestingly, the GDS similarly had higher agreements with worsening cognitive status, despite it being designed for older adults without cognitive impairment. The reason for this is unclear.

Lastly, while others have noted a decrease in GDS and NPIQ agreement with worsening cognition^27^,the NPIQ threshold of >1 and =0 was more intransient to this phenomenon. However, a widening asymmetry in positive predictive agreements between NPIQ and GDS measures was noted with worsening cognition; specifically, the informant-rated NPIQ indicated depression was present more often while the self-report GDS tended to indicate depression absence.

### Affective NPS frequency by etiology

The NACC dataset captures a comparatively large number of participants with different dementia etiologies, making it an important resource for assessing affective NPS frequency by etiology. While greater apathy frequency in FTD was expected based on the diagnostic criteria for behavioral variant FTD^55^, the increase in anxiety and depression by ∼10% across metrics for LBD compared to other etiologies is not similarly explained by being a core criteria of LBD. Whether Lewy Bodies or pathological alpha-synuclein, even at subsyndromal levels, might be especially associated with affective NPS in dementia is an intriguing hypothesis^56–58^. It was also surprising that VCI, which has been noted to have higher rates of depression in some cohorts^59,60^, had similar rates of depression and anxiety to AD while having lower rates of apathy. Large cohort studies with broad inclusion criteria will be better able to capture the true rates of these affective NPS to confirm these patterns.

### Limitations

While the greatest strength of this study is the large number of participants and clinical assessment, multiple limitations should be noted. For instance, there is heterogeneity in recruitment strategies and differences in NPS metric determination that were likely across research sites. Again, with this dataset being a case series, any significant associations with clinical variables may best be considered hypotheses to be investigated in large, prospective cohorts. The lack of a gold standard for NPS diagnosis is a notable limitation when interpreting NPIQ thresholds and NPS frequency. Still, we suggest that this highlights the opportunity for more tailored diagnostic schemes for NPS in dementia. It was notable that only one validated scale for a single syndrome, the GDS, was included in this dataset, and the NPIQ is broad but less rigorous than other forms of the NPI. Other rating scales such as the Cornell Scale for Depression, Rating for Anxiety in Dementia, and Dimensional Apathy Scale would be more appropriate to compare to the NPIQ and serve as future opportunities for studies understanding how NPS in dementia are determined. This study did not use pathological or biomarker confirmation of these etiologies, instead relying on the diagnoses of clinical syndromes. However, as this case series and many clinicians are documenting these biomarkers more frequently, future opportunities to restrict the analysis to those with biomarkers or neuropathologically confirmed etiologies will be important to pursue.

## Acknowledgements

We gratefully acknowledge the participants, their study partners and families whose contributions make this research possible

## Author Contributions

Daniel W. Fisher (Conceptualization, Formal Analysis, Funding Acquisition, Investigation, Methodology, Project Administration, Software, Validation, Visualization, Writing – Original Draft, Reviewing, and Editing), Ronak Mehta (Formal Analysis, Investigation, Methodology, Software, Validation, Writing – Original Draft, Reviewing, and Editing), Christopher B. Morrow (Funding Acquisition, Writing – Reviewing and Editing), Kathleen F. Kerr (Supervision, Writing – Reviewing and Editing), Suman Jayadev (Supervision, Writing – Reviewing and Editing), Kimiko Domoto-Reilly (Supervision, Writing – Reviewing and Editing), Michael J. Schrift (Funding Acquisition, Supervision, Writing – Reviewing and Editing), and Martin Darvas (Conceptualization, Funding Acquisition, Supervision, Writing – Reviewing and Editing)

## Ethical Considerations and Consent

All participants (and their study partners/co-participants) provided written informed consent under protocols approved by each ADRC’s Institutional Review Board.

## Consent for Publication

Not applicable

## Data Availability

The raw data for the Uniform Data Set from NACC that supports these findings are openly available after appropriate request at https://naccdata.org/requesting-data/nacc-data. The analyzed data supporting the findings of this study are available on request from the corresponding author.

