## Supplementary Figures 1-6 for "Affective Neuropsychiatric Symptom Metrics in the National Alzheimer’s Coordinating Center Dataset"

**A**

| | Measure 1 | Measure 2 | PPA: Metric 2 given Metric 1 | PPA: Metric 1 given Metric 2 | NPA: Metric 2 given Metric 1 | NPA: Metric 1 given Metric 2 | Jaccard + | Jaccard - |
|---|---|---|---|---|---|---|---|---|
| Clinical Gestalt | Clinician-Rated | EMR/Report | 0.87 | 0.55 | 0.84 | 0.97 | 0.51 | 0.81 |
| | Consensus-Rated | EMR/Report | 0.88 | 0.51 | 0.83 | 0.97 | 0.47 | 0.81 |
| | Consensus-Rated | Clinician-Rated | 0.67 | 0.60 | 0.92 | 0.94 | 0.46 | 0.87 |
| GDS | GDS | EMR/Report | 0.72 | 0.23 | 0.76 | 0.96 | 0.21 | 0.74 |
| | GDS | Clinician-Rated | 0.55 | 0.28 | 0.88 | 0.96 | 0.23 | 0.84 |
| | GDS | Consensus-Rated | 0.54 | 0.30 | 0.89 | 0.96 | 0.24 | 0.85 |
| NPIQ > 0 | GDS | NPIQ > 0 | 0.60 | 0.22 | 0.81 | 0.96 | 0.19 | 0.78 |
| | NPIQ > 0 | EMR/Report | 0.64 | 0.53 | 0.81 | 0.87 | 0.41 | 0.73 |
| | NPIQ > 0 | Clinician-Rated | 0.54 | 0.71 | 0.93 | 0.87 | 0.44 | 0.82 |
| | NPIQ > 0 | Consensus-Rated | 0.39 | 0.59 | 0.92 | 0.83 | 0.31 | 0.78 |
| NPIQ > 1 | GDS | NPIQ > 1 | 0.30 | 0.33 | 0.95 | 0.94 | 0.18 | 0.89 |
| | NPIQ > 1 | EMR/Report | 0.74 | 0.21 | 0.74 | 0.97 | 0.19 | 0.73 |
| | NPIQ > 1 | Clinician-Rated | 0.65 | 0.28 | 0.86 | 0.97 | 0.25 | 0.84 |
| | NPIQ > 1 | Consensus-Rated | 0.48 | 0.24 | 0.87 | 0.95 | 0.19 | 0.84 |
| NPIQ > 2 | GDS | NPIQ > 2 | 0.06 | 0.45 | 0.99 | 0.92 | 0.06 | 0.92 |
| | NPIQ > 2 | EMR/Report | 0.82 | 0.04 | 0.71 | 1.00 | 0.04 | 0.71 |
| | NPIQ > 2 | Clinician-Rated | 0.73 | 0.05 | 0.83 | 1.00 | 0.05 | 0.83 |
| | NPIQ > 2 | Consensus-Rated | 0.57 | 0.05 | 0.85 | 0.99 | 0.04 | 0.85 |
| NPIQ >1 and <0 | GDS | NPIQ > 1 and < 0 | 0.42 | 0.33 | 0.94 | 0.96 | 0.23 | 0.90 |
| | NPIQ > 1 and < 0 | EMR/Report | 0.74 | 0.30 | 0.81 | 0.97 | 0.28 | 0.79 |
| | NPIQ > 1 and < 0 | Clinician-Rated | 0.65 | 0.49 | 0.93 | 0.96 | 0.39 | 0.90 |
| | NPIQ > 1 and < 0 | Consensus-Rated | 0.48 | 0.37 | 0.92 | 0.95 | 0.26 | 0.87 |

**B**

| | Measure 1 | Measure 2 | PPA: Metric 2 given Metric 1 | PPA: Metric 1 given Metric 2 | NPA: Metric 2 given Metric 1 | NPA: Metric 1 given Metric 2 | Jaccard + | Jaccard - |
|---|---|---|---|---|---|---|---|---|
| Clinical Gestalt | Clinician-Rated | EMR/Report | 0.57 | 0.52 | 0.88 | 0.90 | 0.38 | 0.81 |
| | Consensus-Rated | EMR/Report | 0.73 | 0.24 | 0.84 | 0.98 | 0.22 | 0.82 |
| | Consensus-Rated | Clinician-Rated | 0.65 | 0.20 | 0.86 | 0.98 | 0.18 | 0.84 |
| NPIQ > 0 | NPIQ > 0 | EMR/Report | 0.42 | 0.54 | 0.88 | 0.82 | 0.31 | 0.73 |
| | NPIQ > 0 | Clinician-Rated | 0.61 | 0.79 | 0.95 | 0.89 | 0.52 | 0.86 |
| | NPIQ > 0 | Consensus-Rated | 0.13 | 0.56 | 0.97 | 0.81 | 0.12 | 0.79 |
| NPIQ > 1 | NPIQ > 1 | EMR/Report | 0.49 | 0.26 | 0.84 | 0.93 | 0.20 | 0.79 |
| | NPIQ > 1 | Clinician-Rated | 0.67 | 0.33 | 0.87 | 0.97 | 0.29 | 0.85 |
| | NPIQ > 1 | Consensus-Rated | 0.16 | 0.26 | 0.96 | 0.93 | 0.11 | 0.89 |
| NPIQ > 2 | NPIQ > 2 | EMR/Report | 0.57 | 0.05 | 0.81 | 0.99 | 0.05 | 0.80 |
| | NPIQ > 2 | Clinician-Rated | 0.72 | 0.06 | 0.84 | 0.99 | 0.06 | 0.83 |
| | NPIQ > 2 | Consensus-Rated | 0.19 | 0.06 | 0.95 | 0.99 | 0.04 | 0.94 |
| NPIQ > 1 and < 0 | NPIQ > 1 and < 0 | EMR/Report | 0.49 | 0.36 | 0.88 | 0.93 | 0.26 | 0.82 |
| | NPIQ > 1 and < 0 | Clinician-Rated | 0.67 | 0.61 | 0.95 | 0.96 | 0.47 | 0.92 |
| | NPIQ > 1 and < 0 | Consensus-Rated | 0.16 | 0.37 | 0.97 | 0.92 | 0.12 | 0.89 |

**C**

| | Measure 1 | Measure 2 | PPA: Metric 2 given Metric 1 | PPA: Metric 1 given Metric 2 | NPA: Metric 2 given Metric 1 | NPA: Metric 1 given Metric 2 | Jaccard + | Jaccard - |
|---|---|---|---|---|---|---|---|---|
| | NPIQ > 0 | Clinician-Rated | 0.67 | 0.74 | 0.94 | 0.92 | 0.55 | 0.87 |
| | NPIQ > 1 | Clinician-Rated | 0.76 | 0.40 | 0.88 | 0.97 | 0.36 | 0.86 |
| | NPIQ > 2 | Clinician-Rated | 0.83 | 0.13 | 0.84 | 0.99 | 0.13 | 0.83 |
| | NPIQ > 1 and < 0 | Clinician-Rated | 0.76 | 0.61 | 0.94 | 0.97 | 0.51 | 0.92 |

**Supplementary Figure 1: NPS agreement measures with directional agreements.** PPA determines the directional agreement when a symptom is present, while NPA determines the directional agreement for when a symptom is absent. Specifically, if these percent agreements represent the ratio of times one metric is positive or negative and the other metric agrees. Jaccard indices detail agreement across two metrics for when a NPS is present (Jaccard+) and when it is absent (Jaccard-), where the ratio depicts the total agreement over the total amount either metric suggests presence or absence. Degree of agreement is depicted as a number of 0 (no agreement) to 1 (total agreement) with increased color intensity for agreement for presence (blue) and agreement for absence (orange). Results of agreement for different NPI thresholds are shown for **A)** Depression, **B)** Anxiety, and **C)** Apathy. Positive Predictive Agreement, PPA; Negative Predictive Agreement, NPA; Neuropsychiatric Symptom, NPS; Geriatric Depression Scale, GDS; Neuropsychiatric Inventory Questionnaire, NPIQ; Electronic Medical Record; EMR.

**A**

| | Measure 1 | Measure 2 | Youden's J-like |
|---|---|---|---|
| GDS by NPIQ | GDS | NPIQ > 0 | -0.03 |
| | GDS | NPIQ > 1 | 0.08 |
| | GDS | NPIQ > 2 | -0.02 |
| | GDS | NPIQ > 1 and < 0 | 0.13 |
| EMR/Report by NPIQ | NPIQ > 0 | EMR/Report | -0.03 |
| | NPIQ > 1 | EMR/Report | 0.08 |
| | NPIQ > 2 | EMR/Report | -0.02 |
| | NPIQ > 1 and < 0 | EMR/Report | 0.07 |
| Clinician-Rated by NPIQ | NPIQ > 0 | Clinician-Rated | 0.26 |
| | NPIQ > 1 | Clinician-Rated | 0.09 |
| | NPIQ > 2 | Clinician-Rated | -0.12 |
| | NPIQ > 1 and < 0 | Clinician-Rated | 0.29 |
| Consensus-Rated by NPIQ | NPIQ > 0 | Consensus-Rated | 0.09 |
| | NPIQ > 1 | Consensus-Rated | 0.08 |
| | NPIQ > 2 | Consensus-Rated | 0.03 |
| | NPIQ > 1 and < 0 | Consensus-Rated | 0.14 |

**B**

| | Measure 1 | Measure 2 | Youden's J-like |
|---|---|---|---|
| EMR/Report | NPIQ > 0 | EMR/Report | 0.04 |
| | NPIQ > 1 | EMR/Report | -0.01 |
| | NPIQ > 2 | EMR/Report | -0.15 |
| | NPIQ > 1 and < 0 | EMR/Report | 0.08 |
| Clinician-Rated | NPIQ > 0 | Clinician-Rated | 0.38 |
| | NPIQ > 1 | Clinician-Rated | 0.13 |
| | NPIQ > 2 | Clinician-Rated | -0.11 |
| | NPIQ > 1 and < 0 | Clinician-Rated | 0.39 |
| Consensus-Rated | NPIQ > 0 | Consensus-Rated | -0.09 |
| | NPIQ > 1 | Consensus-Rated | 0.00 |
| | NPIQ > 2 | Consensus-Rated | -0.01 |
| | NPIQ > 1 and < 0 | Consensus-Rated | 0.02 |

**C**

| Measure 1 | Measure 2 | Youden's J-like |
|---|---|---|
| NPIQ > 0 | Clinician-Rated | 0.42 |
| NPIQ > 1 | Clinician-Rated | 0.21 |
| NPIQ > 2 | Clinician-Rated | -0.04 |
| NPIQ > 1 and < 0 | Clinician-Rated | 0.43 |

**Supplementary Figure 2: Youden's J-like Statistics.** To get a sense of the balance of agreement amongst the presence and absence of NPS, a Youden's J-like statistic was calculated from the Jaccard+ and Jaccard- values for **A)** Depression, **B)** Anxiety, and **C)** Apathy. Neuropsychiatric Inventory Questionnaire, NPIQ; Electronic Medical Record; EMR.

**A**

| | Metric 1 | Metric 2 | Cohen's Kappa | CI Lower | CI Upper |
|---|---|---|---|---|---|
| GDS by NPIQ | GDS | NPIQ > 0 | 0.23 | 0.22 | 0.23 |
| | GDS | NPIQ > 1 | 0.26 | 0.25 | 0.26 |
| | GDS | NPIQ > 2 | 0.09 | 0.09 | 0.10 |
| | GDS | NPIQ > 1 and < 0 | 0.32 | 0.31 | 0.33 |
| EMR/Report by NPIQ | NPIQ > 0 | EMR/Report | 0.42 | 0.42 | 0.43 |
| | NPIQ > 1 | EMR/Report | 0.22 | 0.22 | 0.23 |
| | NPIQ > 2 | EMR/Report | 0.05 | 0.05 | 0.05 |
| | NPIQ > 1 and < 0 | EMR/Report | 0.34 | 0.33 | 0.35 |
| Clinician-Rated by NPIQ | NPIQ > 0 | Clinician-Rated | 0.51 | 0.51 | 0.52 |
| | NPIQ > 1 | Clinician-Rated | 0.32 | 0.32 | 0.33 |
| | NPIQ > 2 | Clinician-Rated | 0.08 | 0.07 | 0.08 |
| | NPIQ > 1 and < 0 | Clinician-Rated | 0.51 | 0.50 | 0.51 |
| Consensus-Rated by NPIQ | NPIQ > 0 | Consensus-Rated | 0.35 | 0.35 | 0.36 |
| | NPIQ > 1 | Consensus-Rated | 0.24 | 0.24 | 0.25 |
| | NPIQ > 2 | Consensus-Rated | 0.06 | 0.06 | 0.07 |
| | NPIQ > 1 and < 0 | Consensus-Rated | 0.35 | 0.34 | 0.36 |

**B**

| | Metric 1 | Metric 2 | Cohen's Kappa | CI Lower | CI Upper |
|---|---|---|---|---|---|
| EMR/Report | NPIQ > 0 | EMR/Report | 0.32 | 0.30 | 0.33 |
| | NPIQ > 1 | EMR/Report | 0.23 | 0.21 | 0.25 |
| | NPIQ > 2 | EMR/Report | 0.06 | 0.05 | 0.08 |
| | NPIQ > 1 and < 0 | EMR/Report | 0.32 | 0.30 | 0.34 |
| Clinician-Rated | NPIQ > 0 | Clinician-Rated | 0.61 | 0.60 | 0.62 |
| | NPIQ > 1 | Clinician-Rated | 0.37 | 0.36 | 0.38 |
| | NPIQ > 2 | Clinician-Rated | 0.09 | 0.08 | 0.10 |
| | NPIQ > 1 and < 0 | Clinician-Rated | 0.60 | 0.59 | 0.61 |
| Consensus-Rated | NPIQ > 0 | Consensus-Rated | 0.14 | 0.13 | 0.15 |
| | NPIQ > 1 | Consensus-Rated | 0.14 | 0.13 | 0.15 |
| | NPIQ > 2 | Consensus-Rated | 0.07 | 0.05 | 0.08 |
| | NPIQ > 1 and < 0 | Consensus-Rated | 0.17 | 0.16 | 0.19 |

**C**

| Metric 1 | Metric 2 | Cohen's Kappa | CI Lower | CI Upper |
|---|---|---|---|---|
| NPIQ > 0 | Clinician-Rated | 0.64 | 0.63 | 0.64 |
| NPIQ > 1 | Clinician-Rated | 0.46 | 0.45 | 0.46 |
| NPIQ > 2 | Clinician-Rated | 0.19 | 0.18 | 0.20 |
| NPIQ > 1 and < 0 | Clinician-Rated | 0.63 | 0.63 | 0.64 |

**D**

| | Metric 1 | Metric 2 | Cohen's Kappa | CI Lower | CI Upper |
|---|---|---|---|---|---|
| Clinical Gestalt | Clinician-Rated | EMR/Report | 0.58 | 0.57 | 0.58 |
| | Consensus-Rated | EMR/Report | 0.55 | 0.54 | 0.55 |
| | Consensus-Rated | Clinician-Rated | 0.56 | 0.55 | 0.56 |
| Objective | GDS | NPIQ > 1 and < 0 | 0.32 | 0.31 | 0.33 |
| Clinical Gestalt - Objective | EMR/Report | GDS | 0.24 | 0.23 | 0.25 |
| | Clinician-Rated | GDS | 0.30 | 0.29 | 0.30 |
| | Consensus-Rated | GDS | 0.31 | 0.30 | 0.32 |
| | EMR/Report | NPIQ > 1 and < 0 | 0.34 | 0.33 | 0.35 |
| | Clinician-Rated | NPIQ > 1 and < 0 | 0.51 | 0.50 | 0.51 |
| | Consensus-Rated | NPIQ > 1 and < 0 | 0.35 | 0.35 | 0.36 |

**E**

| | Metric 1 | Metric 2 | Cohen's Kappa | CI Lower | CI Upper |
|---|---|---|---|---|---|
| Clinical Gestalt | Clinician-Rated | EMR/Report | 0.44 | 0.42 | 0.46 |
| | Consensus-Rated | EMR/Report | 0.29 | 0.28 | 0.31 |
| | Consensus-Rated | Clinician-Rated | 0.25 | 0.24 | 0.26 |
| Clinical Gestalt - Objective | EMR/Report | NPIQ > 1 and < 0 | 0.32 | 0.30 | 0.34 |
| | Clinician-Rated | NPIQ > 1 and < 0 | 0.60 | 0.59 | 0.61 |
| | Consensus-Rated | NPIQ > 1 and < 0 | 0.17 | 0.16 | 0.19 |

**F**

| Approaches | Cohen's Kappa | CI Lower | CI Upper |
|---|---|---|---|
| **GDS-Gold Standard*** | 0.39 | 0.37 | 0.41 |
| GDS-NPIQ >1 and <0 | 0.32 | 0.31 | 0.33 |
| Panel-GDS | 0.31 | 0.3 | 0.32 |
| Clinician-GDS | 0.3 | 0.29 | 0.3 |
| GDS-NPIQ > 1 | 0.26 | 0.25 | 0.26 |
| GDS-EMR | 0.24 | 0.23 | 0.25 |
| GDS-NPIQ > 0 | 0.23 | 0.22 | 0.23 |
| GDS-NPIQ > 2 | 0.09 | 0.09 | 0.1 |

**Supplementary Figure 3: Cohen's Kappa Agreement Measures.** Cohen's Kappa with 95% Confidence Intervals (CI) shown for **A)** Depression, **B)** Anxiety, and **C)** Apathy by NPIQ cut-offs. Comparison of all metric with NPIQ cut off >1 and =0 shown for **D)** Depression and **E)** Anxiety. **F)** Gold-standard clinician diagnosis of Major Depressive Disorder was compared to GDS performance (bolded and starred) and compared to depression metrics in the UDS. Neuropsychiatric Symptom, NPS; Geriatric Depression Scale, GDS; Neuropsychiatric Inventory Questionnaire, NPIQ; Electronic Medical Record; EMR.

| NPIQ > 0 | NPIQ > 1 | NPIQ > 2 | NPIQ > 1 and < 0 |
|---|---|---|---|

**A**

| Metrics | Jaccard + |
|---|---|
| Clinician - NPIQ > 0 | 0.44 |
| EMR/Report - NPIQ > 0 | 0.41 |
| Clinician - NPIQ > 1 and < 0 | 0.39 |
| Consensus - NPIQ > 0 | 0.31 |
| EMR/Report - Clinician - NPIQ > 0 | 0.28 |
| EMR/Report - NPIQ > 1 and < 0 | 0.28 |
| Consensus - NPIQ > 1 and < 0 | 0.26 |
| Clinician - NPIQ > 1 | 0.25 |
| EMR/Report - Consensus - NPIQ > 0 | 0.24 |
| Clinician - Consensus - NPIQ > 0 | 0.23 |
| NPIQ > 1 and < 0 - GDS | 0.23 |
| EMR/Report - Clinician - ,NPIQ > 1 and < 0 | 0.20 |
| Clinician - Consensus - NPIQ > 1 and < 0 | 0.19 |
| EMR/Report - NPIQ > 1 | 0.19 |
| EMR/Report - Clinician - Consensus - NPIQ > 0 | 0.19 |
| Consensus - NPIQ > 1 | 0.19 |
| NPIQ > 0 - GDS | 0.19 |
| NPIQ > 1 - GDS | 0.18 |
| EMR/Report - Consensus - NPIQ > 1 and < 0 | 0.17 |
| EMR/Report - Clinician - NPIQ > 1 | 0.14 |
| EMR/Report - Clinician - Consensus - NPIQ > 1 and < 0 | 0.14 |
| Clinician - Consensus - NPIQ > 1 | 0.13 |
| Clinician - NPIQ > 1 and < 0 - GDS | 0.13 |
| Clinician - NPIQ > 0 - GDS | 0.12 |
| EMR/Report - Consensus - NPIQ > 1 | 0.12 |
| EMR/Report - NPIQ > 0 - GDS | 0.11 |
| Consensus - NPIQ > 1 and < 0 - GDS | 0.10 |
| EMR/Report - NPIQ > 1 and < 0,GDS | 0.10 |
| Consensus - NPIQ > 0 - GDS | 0.10 |
| EMR/Report - Clinician - Consensus - NPIQ > 1 | 0.10 |
| EMR/Report - Clinician - NPIQ > 0 - GDS | 0.09 |
| Clinician - NPIQ > 1 - GDS | 0.08 |
| EMR/Report - Consensus - NPIQ > 0 - GDS | 0.08 |
| Clinician - Consensus - NPIQ > 1 and < 0 - GDS | 0.08 |
| Clinician - Consensus - NPIQ > 0 - GDS | 0.08 |
| EMR/Report - Clinician - NPIQ > 1 and < 0 - GDS | 0.08 |
| Consensus - NPIQ > 1 - GDS | 0.08 |
| EMR/Report - Consensus - NPIQ > 1 and < 0 - GDS | 0.07 |
| EMR/Report - NPIQ > 1 - GDS | 0.07 |
| EMR/Report - Clinician - Consensus - NPIQ > 0 - GDS | 0.07 |
| EMR/Report - Clinician - Consensus - NPIQ > 1 and < 0 - GDS | 0.06 |
| NPIQ > 2 - GDS | 0.06 |
| EMR/Report,Clinician - NPIQ > 1 - GDS | 0.05 |
| Clinician - Consensus - NPIQ > 1 - GDS | 0.05 |
| EMR/Report - Consensus - NPIQ > 1 - GDS | 0.05 |
| Clinician - NPIQ > 2 | 0.05 |
| Consensus - NPIQ > 2 | 0.04 |
| EMR/Report - Clinician - Consensus - NPIQ > 1 - GDS | 0.04 |
| EMR/Report - NPIQ > 2 | 0.04 |
| EMR/Report - Clinician - NPIQ > 2 | 0.03 |
| Clinician - Consensus - NPIQ > 2 | 0.03 |
| EMR/Report - Consensus - NPIQ > 2 | 0.02 |
| Clinician - NPIQ > 2 - GDS | 0.02 |
| EMR/Report - Clinician - Consensus - NPIQ > 2 | 0.02 |
| Consensus - NPIQ > 2 - GDS | 0.02 |
| EMR/Report - NPIQ > 2 - GDS | 0.02 |
| Clinician - Consensus - NPIQ > 2 - GDS | 0.01 |
| EMR/Report - Clinician - NPIQ > 2 - GDS | 0.01 |
| EMR/Report - Consensus - NPIQ > 2 - GDS | 0.01 |
| EMR/Report - Clinician - Consensus - NPIQ > 2 - GDS | 0.01 |

**C**

| Metrics | Jaccard - |
|---|---|
| NPIQ > 2 - GDS | 0.92 |
| NPIQ > 1 and < 0 - GDS | 0.90 |
| Clinician - NPIQ > 1 and < 0 | 0.90 |
| NPIQ > 1 - GDS | 0.89 |
| Consensus - NPIQ > 1 and < 0 | 0.87 |
| Clinician - NPIQ > 1 and < 0 - GDS | 0.85 |
| Consensus - NPIQ > 2 | 0.85 |
| Clinician - Consensus - NPIQ > 1 and < 0 | 0.84 |
| Clinician - NPIQ > 1 | 0.84 |
| Consensus - NPIQ > 1 | 0.84 |
| Consensus - NPIQ > 1 and < 0 - GDS | 0.83 |
| Clinician - NPIQ > 2 | 0.83 |
| Clinician - NPIQ > 0 | 0.82 |
| Consensus - NPIQ > 2 - GDS | 0.81 |
| Clinician - Consensus - NPIQ > 1 and < 0 - GDS | 0.81 |
| Clinician - NPIQ > 2 - GDS | 0.81 |
| Consensus - NPIQ > 1 - GDS | 0.80 |
| Clinician - NPIQ > 1 - GDS | 0.80 |
| EMR/Report - NPIQ > 1 and < 0 | 0.79 |
| NPIQ > 0 - GDS | 0.78 |
| Clinician - Consensus - NPIQ > 1 | 0.78 |
| Clinician - Consensus - NPIQ > 2 | 0.78 |
| Consensus - NPIQ > 0 | 0.78 |
| EMR/Report - Clinician - NPIQ > 1 and < 0 | 0.77 |
| Clinician - Consensus - NPIQ > 2 - GDS | 0.76 |
| EMR/Report - Consensus - NPIQ > 1 and < 0 | 0.76 |
| Clinician - Consensus - NPIQ > 1 - GDS | 0.76 |
| EMR/Report - NPIQ > 1 and < 0 - GDS | 0.75 |
| EMR/Report - Clinician - Consensus - NPIQ > 1 and < 0 | 0.74 |
| Clinician - Consensus - NPIQ > 0 | 0.74 |
| Clinician - NPIQ > 0 - GDS | 0.73 |
| EMR/Report - Clinician - NPIQ > 1 and < 0 - GDS | 0.73 |
| EMR/Report - Consensus - NPIQ > 1 and < 0 - GDS | 0.73 |
| EMR/Report - NPIQ > 0 | 0.73 |
| EMR/Report - NPIQ > 1 | 0.73 |
| EMR/Report - Clinician - Consensus - NPIQ > 1 and < 0 - GDS | 0.72 |
| Consensus - NPIQ > 0 - GDS | 0.72 |
| EMR/Report - NPIQ > 2 | 0.71 |
| Clinician - Consensus - NPIQ > 0 - GDS | 0.70 |
| EMR/Report - Clinician - NPIQ > 1 | 0.70 |
| EMR/Report - NPIQ > 2 - GDS | 0.69 |
| EMR/Report - Consensus - NPIQ > 1 | 0.69 |
| EMR/Report - NPIQ > 1 - GDS | 0.69 |
| EMR/Report - Consensus - NPIQ > 2 | 0.69 |
| EMR/Report - Clinician - NPIQ > 2 | 0.68 |
| EMR/Report - Clinician - NPIQ > 0 | 0.68 |
| EMR/Report - Consensus - NPIQ > 2 - GDS | 0.68 |
| EMR/Report - Clinician - Consensus - NPIQ > 1 | 0.67 |
| EMR/Report - Clinician - NPIQ > 2 - GDS | 0.67 |
| EMR/Report - Consensus - NPIQ > 1 - GDS | 0.67 |
| EMR/Report - Clinician - NPIQ > 1 - GDS | 0.67 |
| EMR/Report - Clinician - Consensus - NPIQ > 2 | 0.67 |
| EMR/Report - Consensus - NPIQ > 0 | 0.67 |
| EMR/Report - Clinician - Consensus - NPIQ > 2 - GDS | 0.66 |
| EMR/Report - Clinician - Consensus - NPIQ > 1 - GDS | 0.66 |
| EMR/Report - Clinician - Consensus - NPIQ > 0 | 0.65 |
| EMR/Report - NPIQ > 0 - GDS | 0.64 |
| EMR/Report - Clinician - NPIQ > 0 - GDS | 0.63 |
| EMR/Report - Consensus - NPIQ > 0 - GDS | 0.62 |
| EMR/Report - Clinician - Consensus - NPIQ > 0 - GDS | 0.61 |

**B**

| Metrics | Jaccard + |
|---|---|
| EMR/Report - Clinician | 0.51 |
| EMR/Report - Consensus | 0.47 |
| Clinician - Consensus | 0.46 |
| Clinician - NPIQ | 0.39 |
| EMR/Report - Clinician - Consensus | 0.33 |
| EMR/Report - NPIQ | 0.28 |
| Consensus - NPIQ | 0.26 |
| Consensus - GDS | 0.24 |
| Clinician - GDS | 0.23 |
| NPIQ - GDS | 0.23 |
| EMR/Report - GDS | 0.21 |
| EMR/Report - Clinician - NPIQ | 0.20 |
| Clinician - Consensus - NPIQ | 0.19 |
| EMR/Report - Consensus - NPIQ | 0.17 |
| Clinician - Consensus - GDS | 0.14 |
| EMR/Report - Consensus - GDS | 0.14 |
| EMR/Report - Clinician - Consensus - NPIQ | 0.14 |
| EMR/Report - Clinician - GDS | 0.14 |
| Clinician - NPIQ - GDS | 0.13 |
| Consensus - NPIQ - GDS | 0.10 |
| EMR/Report - Clinician - Consensus - GDS | 0.10 |
| EMR/Report - NPIQ - GDS | 0.10 |
| Clinician - Consensus - NPIQ - GDS | 0.08 |
| EMR/Report - Clinician - NPIQ - GDS | 0.08 |
| EMR/Report - Consensus - NPIQ - GDS | 0.07 |
| EMR/Report - Clinician - Consensus - NPIQ - GDS | 0.06 |

**D**

| Metrics | Jaccard - |
|---|---|
| NPIQ - GDS | 0.90 |
| Clinician - NPIQ | 0.90 |
| Consensus - NPIQ | 0.87 |
| Clinician - Consensus | 0.87 |
| Consensus - GDS | 0.85 |
| Clinician - NPIQ - GDS | 0.85 |
| Clinician - GDS | 0.84 |
| Clinician - Consensus - NPIQ | 0.84 |
| Consensus - NPIQ - GDS | 0.83 |
| EMR/Report - Clinician | 0.81 |
| Clinician - Consensus - NPIQ - GDS | 0.81 |
| EMR/Report - Consensus | 0.81 |
| EMR/Report - NPIQ | 0.79 |
| Clinician - Consensus - GDS | 0.79 |
| EMR/Report - Clinician - NPIQ | 0.77 |
| EMR/Report - Consensus - NPIQ | 0.76 |
| EMR/Report - Clinician - Consensus | 0.75 |
| EMR/Report - NPIQ - GDS | 0.75 |
| EMR/Report - Clinician - Consensus - NPIQ | 0.74 |
| EMR/Report - GDS | 0.74 |
| EMR/Report - Clinician - NPIQ - GDS | 0.73 |
| EMR/Report - Consensus - NPIQ - GDS | 0.73 |
| EMR/Report - Clinician - Consensus - NPIQ - GDS | 0.72 |
| EMR/Report - Consensus - GDS | 0.71 |
| EMR/Report - Clinician - GDS | 0.71 |
| EMR/Report - Clinician - Consensus - GDS | 0.69 |

**Supplementary Figure 4: Joint Agreement for Depression metrics.** Jaccard indices detail agreement across multiple metrics for when a NPS is present (Jaccard+) and when it is absent (Jaccard-), where the ratio depicts the total agreement over the total amount when any metric suggests presence or absence. Degree of agreement is depicted as a number of 0 (no agreement) to 1 (total agreement) with increased color intensity for agreement for presence (blue) and agreement for absence (orange). Results of agreement for **A)** presence among all NPIQ threshold, **B)** presence with only NPIQ > 1 and <0, **C)** absence among all NPIQ thresholds, and **D)** absence with only NPIQ > 1 and < 0. In A and C, each NPIQ threshold is colored: >0 is blue, >1 is yellow, >2 is orange, and >3 is purple. In B and D, a metric combination highlighted in yellow depicts and instance when a combination of >2 metrics has better joint agreement than a pair of metrics. Neuropsychiatric Symptom, NPS; Geriatric Depression Scale, GDS; Neuropsychiatric Inventory Questionnaire, NPIQ; Electronic Medical Record; EMR.

NPIQ > 0 | NPIQ > 1 | NPIQ > 2 | NPIQ > 1 and < 0

**A**

| Metrics | Jaccard + |
|---|---|
| Clinician - NPIQ > 0 | 0.52 |
| Clinician - NPIQ > 1 and < 0 | 0.47 |
| EMR/Report - NPIQ > 0 | 0.31 |
| Clinician - NPIQ > 1 | 0.29 |
| EMR/Report - NPIQ > 1 and < 0 | 0.26 |
| EMR/Report - Clinician - NPIQ > 0 | 0.21 |
| EMR/Report - NPIQ > 1 | 0.20 |
| EMR/Report - Clinician - NPIQ > 1 and < 0 | 0.18 |
| EMR/Report - Clinician - NPIQ > 1 | 0.13 |
| Consensus - NPIQ > 1 and < 0 | 0.12 |
| Consensus - NPIQ > 0 | 0.12 |
| Consensus - NPIQ > 1 | 0.11 |
| Clinician - Consensus - NPIQ > 0 | 0.09 |
| EMR/Report - Consensus - NPIQ > 0 | 0.09 |
| Clinician - Consensus - NPIQ > 1 and < 0 | 0.09 |
| EMR/Report - Consensus - NPIQ > 1 and < 0 | 0.08 |
| EMR/Report - Clinician - Consensus - NPIQ > 0 | 0.07 |
| EMR/Report - Clinician - Consensus - NPIQ > 1 and < 0 | 0.07 |
| Clinician - NPIQ > 2 | 0.06 |
| EMR/Report - Consensus - NPIQ > 1 | 0.06 |
| Clinician - Consensus - NPIQ > 1 | 0.06 |
| EMR/Report - NPIQ > 2 | 0.05 |
| EMR/Report - Clinician - Consensus - NPIQ > 1 | 0.05 |
| Consensus - NPIQ > 2 | 0.04 |
| EMR/Report - Clinician - NPIQ > 2 | 0.03 |
| EMR/Report - Consensus - NPIQ > 2 | 0.02 |
| Clinician - Consensus - NPIQ > 2 | 0.01 |
| EMR/Report - Clinician - Consensus - NPIQ > 2 | 0.01 |

**C**

| Metrics | Jaccard - |
|---|---|
| Consensus - NPIQ > 2 | 0.94 |
| Clinician - NPIQ > 1 and < 0 | 0.92 |
| Consensus - NPIQ > 1 and < 0 | 0.89 |
| Consensus - NPIQ > 1 | 0.89 |
| Clinician - NPIQ > 0 | 0.86 |
| Clinician - Consensus - NPIQ > 1 and < 0 | 0.85 |
| Clinician - NPIQ > 1 | 0.85 |
| Clinician - NPIQ > 2 | 0.83 |
| EMR/Report - NPIQ > 1 and < 0 | 0.82 |
| Clinician - Consensus - NPIQ > 2 | 0.81 |
| EMR/Report - NPIQ > 2 | 0.80 |
| Clinician - Consensus - NPIQ > 1 | 0.79 |
| EMR/Report - NPIQ > 1 | 0.79 |
| Consensus - NPIQ > 0 | 0.79 |
| EMR/Report - Clinician - NPIQ > 1 and < 0 | 0.78 |
| EMR/Report - Consensus - NPIQ > 2 | 0.78 |
| EMR/Report - Consensus - NPIQ > 1 and < 0 | 0.78 |
| EMR/Report - Clinician - Consensus - NPIQ > 1 and < 0 | 0.75 |
| EMR/Report - Consensus - NPIQ > 1 | 0.75 |
| Clinician - Consensus - NPIQ > 0 | 0.74 |
| EMR/Report - NPIQ > 0 | 0.73 |
| EMR/Report - Clinician - NPIQ > 2 | 0.73 |
| EMR/Report - Clinician - NPIQ > 1 | 0.72 |
| EMR/Report - Clinician - Consensus - NPIQ > 2 | 0.72 |
| EMR/Report - Clinician - Consensus - NPIQ > 1 | 0.70 |
| EMR/Report - Clinician - NPIQ > 0 | 0.69 |
| EMR/Report - Consensus - NPIQ > 0 | 0.67 |
| EMR/Report - Clinician - Consensus - NPIQ > 0 | 0.65 |

**B**

| Metrics | Jaccard + |
|---|---|
| Clinician - NPIQ | 0.47 |
| EMR/Report - Clinician | 0.38 |
| EMR/Report - NPIQ | 0.26 |
| EMR/Report - Consensus | 0.22 |
| Clinician - Consensus | 0.18 |
| EMR/Report - Clinician - NPIQ | 0.18 |
| Consensus - NPIQ | 0.12 |
| EMR/Report - Clinician - Consensus | 0.12 |
| Clinician - Consensus - NPIQ | 0.09 |
| EMR/Report - Consensus - NPIQ | 0.08 |
| EMR/Report - Clinician - Consensus - NPIQ | 0.07 |

**D**

| Metrics | Jaccard - |
|---|---|
| Clinician - NPIQ | 0.92 |
| Consensus - NPIQ | 0.89 |
| Clinician - Consensus - NPIQ | 0.85 |
| Clinician - Consensus | 0.84 |
| EMR/Report - Consensus | 0.82 |
| EMR/Report - NPIQ | 0.82 |
| EMR/Report - Clinician | 0.81 |
| EMR/Report - Clinician - NPIQ | 0.78 |
| EMR/Report - Consensus - NPIQ | 0.78 |
| EMR/Report - Clinician - Consensus - NPIQ | 0.75 |
| EMR/Report - Clinician - Consensus | 0.74 |

**Supplementary Figure 5: Joint Agreement for Anxiety metrics.** Jaccard indices detail agreement across multiple metrics for when a NPS is present (Jaccard+) and when it is absent (Jaccard-), where the ratio depicts the total agreement over the total amount when any metric suggests presence or absence. Degree of agreement is depicted as a number of 0 (no agreement) to 1 (total agreement) with increased color intensity for agreement for presence (blue) and agreement for absence (orange). Results of agreement for **A)** presence among all NPIQ threshold, **B)** presence with only NPIQ > 1 and <0, **C)** absence among all NPIQ thresholds, and **D)** absence with only NPIQ > 1 and < 0. In A and C, each NPIQ threshold is colored: >0 is blue, >1 is yellow, >2 is orange, and >3 is purple. In B and D, a metric combination highlighted in yellow depicts and instance when a combination of >2 metrics has better joint agreement than a pair of metrics. Neuropsychiatric Symptom, NPS; Geriatric Depression Scale, GDS; Neuropsychiatric Inventory Questionnaire, NPIQ; Electronic Medical Record; EMR.

**A**

| | Metric 1 | Metric 2 | Cognitively Normal (CN) | | | Mild Cognitive Impairment (MCI) | | | Dementia | | |
|---|---|---|---|---|---|---|---|---|---|---|---|
| | | | Kappa | CI_lower | CI_upper | Kappa | CI_lower | CI_upper | Kappa | CI_lower | CI_upper |
| Clinical Gestalt | Clinician-Rated | EMR/Report | 0.16 | 0.15 | 0.17 | 0.64 | 0.63 | 0.65 | 0.69 | 0.68 | 0.7 |
| | Consensus-Rated | EMR/Report | 0.51 | 0.5 | 0.53 | 0.57 | 0.56 | 0.59 | 0.5 | 0.49 | 0.51 |
| | Consensus-Rated | Clinician-Rated | 0.3 | 0.29 | 0.31 | 0.62 | 0.61 | 0.63 | 0.58 | 0.57 | 0.58 |
| Objective | GDS | NPIQ | 0.24 | 0.23 | 0.26 | 0.32 | 0.3 | 0.34 | 0.3 | 0.28 | 0.31 |
| Clinical Gestalt - Objective | GDS | EMR/Report | 0.21 | 0.2 | 0.22 | 0.26 | 0.25 | 0.27 | 0.19 | 0.18 | 0.2 |
| | GDS | Clinician-Rated | 0.22 | 0.21 | 0.24 | 0.29 | 0.28 | 0.3 | 0.24 | 0.23 | 0.25 |
| | GDS | Consensus-Rated | 0.28 | 0.27 | 0.29 | 0.32 | 0.31 | 0.33 | 0.27 | 0.25 | 0.28 |
| | NPIQ | EMR/Report | 0.19 | 0.18 | 0.21 | 0.34 | 0.32 | 0.36 | 0.37 | 0.36 | 0.38 |
| | NPIQ | Clinician-Rated | 0.31 | 0.29 | 0.33 | 0.46 | 0.45 | 0.48 | 0.52 | 0.51 | 0.53 |
| | NPIQ | Consensus-Rated | 0.23 | 0.22 | 0.25 | 0.35 | 0.34 | 0.37 | 0.36 | 0.35 | 0.38 |

**B**

| | Metric 1 | Metric 2 | Cognitively Normal (CN) | | | Mild Cognitive Impairment (MCI) | | | Dementia | | |
|---|---|---|---|---|---|---|---|---|---|---|---|
| | | | Kappa | CI_lower | CI_upper | Kappa | CI_lower | CI_upper | Kappa | CI_lower | CI_upper |
| Clinical Gestalt | Clinician-Rated | EMR/Report | 0.38 | 0.34 | 0.41 | 0.43 | 0.39 | 0.46 | 0.43 | 0.40 | 0.46 |
| | Consensus-Rated | EMR/Report | 0.28 | 0.25 | 0.32 | 0.32 | 0.28 | 0.35 | 0.24 | 0.21 | 0.27 |
| | Consensus-Rated | Clinician-Rated | 0.29 | 0.28 | 0.31 | 0.3 | 0.28 | 0.32 | 0.15 | 0.14 | 0.16 |
| Clinical Gestalt - Objective | NPIQ | EMR/Report | 0.15 | 0.12 | 0.19 | 0.32 | 0.28 | 0.37 | 0.37 | 0.34 | 0.41 |
| | NPIQ | Clinician-Rated | 0.39 | 0.36 | 0.42 | 0.5 | 0.48 | 0.53 | 0.66 | 0.65 | 0.68 |
| | NPIQ | Consensus-Rated | 0.15 | 0.13 | 0.18 | 0.23 | 0.2 | 0.26 | 0.13 | 0.12 | 0.15 |

**C**

| Metric 1 | Metric 2 | Cognitively Normal (CN) | | | Mild Cognitive Impairment (MCI) | | | Dementia | | |
|---|---|---|---|---|---|---|---|---|---|---|
| | | Kappa | CI_lower | CI_upper | Kappa | CI_lower | CI_upper | Kappa | CI_lower | CI_upper |
| NPIQ | Clinician-Rated | 0.29 | 0.26 | 0.33 | 0.43 | 0.41 | 0.45 | 0.57 | 0.56 | 0.57 |

**Supplementary Figure 6: Cohen's Kappa measures across cognitive statuses.** Cohen's Kappa for agreement for different NPS metrics are shown across different cognitive statuses for **A)** Depression, **B)** Anxiety, and **C)** Apathy. Neuropsychiatric Symptom, NPS; Geriatric Depression Scale, GDS; Neuropsychiatric Inventory Questionnaire, NPIQ; Electronic Medical Record; EMR.
