## Supplementary Table 1 for "Affective Neuropsychiatric Symptom Metrics in the National Alzheimer’s Coordinating Center Dataset"

**Continuous Variables**

| | Mean | | | | | Standard Deviation | | | | |
|---|---|---|---|---|---|---|---|---|---|---|
| **Clinical Variable** | **All** | **Cognitively Normal** | **Mild Cognitive Impairment** | **Dementia** | **Impaired-not-MCI** | **All** | **Cognitively Normal** | **Mild Cognitive Impairment** | **Dementia** | **Impaired-not-MCI** |
| **Age at study visit** | 76.1 | 75.4 | 76.8 | 77.0 | 75.6 | 8.4 | 8.1 | 8.3 | 8.8 | 8.0 |
| **Body Mass Index** | 27.0 | 27.3 | 27.0 | 26.4 | 27.5 | 5.2 | 5.3 | 5.2 | 4.8 | 5.3 |
| **CDR-Sum of Boxes** | 2.8 | 0.1 | 1.4 | 8.2 | 0.9 | 4.5 | 0.3 | 1.2 | 5.1 | 1.2 |
| **Cognitive Score** | 21.9 | 26.0 | 22.4 | 13.2 | 24.2 | 7.0 | 2.8 | 3.9 | 6.9 | 3.7 |
| **Diastolic Blood Pressure** | 74.5 | 74.5 | 74.6 | 74.3 | 75.5 | 10.4 | 10.2 | 10.5 | 10.7 | 10.6 |
| **Education Level** | 15.6 | 16.0 | 15.5 | 14.9 | 15.3 | 3.3 | 2.9 | 3.3 | 3.6 | 3.5 |
| **Heart Rate** | 68.0 | 68.1 | 67.7 | 68.3 | 68.1 | 10.8 | 10.6 | 10.9 | 11.2 | 10.8 |
| **Mean Arterial Pressure** | 94.4 | 94.2 | 94.9 | 94.3 | 95.5 | 11.4 | 11.1 | 11.5 | 11.7 | 11.4 |
| **MMSE** | 25.5 | 28.9 | 27.0 | 19.2 | 28.0 | 6.0 | 1.5 | 2.5 | 7.0 | 2.2 |
| **MOCA** | 23.4 | 26.4 | 22.5 | 15.1 | 24.6 | 5.7 | 2.7 | 3.6 | 6.1 | 3.4 |
| **Number of Medications** | 6.9 | 6.7 | 7.1 | 7.1 | 7.2 | 4.2 | 4.2 | 4.3 | 4.1 | 4.3 |
| **Systolic Blood Presure** | 134.2 | 133.5 | 135.6 | 134.2 | 135.5 | 18.6 | 18.1 | 18.8 | 19.3 | 18.2 |

**Categorical Variables**

| | | All | | Cognitively Normal | | Mild Cognitive Impairment | | Dementia | | Impaired-not-MCI | |
|---|---|---|---|---|---|---|---|---|---|---|---|
| **Clinical Variable** | **Value** | **Count** | **Frequency** | **Count** | **Frequency** | **Count** | **Frequency** | **Count** | **Frequency** | **Count** | **Frequency** |
| **Atrial Fibrillation** | **Absent** | 74343 | 91.3 | 40464 | 91.9 | 13160 | 90.1 | 17606 | 91.1 | 3113 | 90.8 |
| | **Present** | 7044 | 8.7 | 3561 | 8.1 | 1447 | 9.9 | 1720 | 8.9 | 316 | 9.2 |
| **Angina** | **Absent** | 79041 | 97.0 | 42898 | 97.1 | 14051 | 96.3 | 18816 | 97.5 | 3276 | 95.5 |
| | **Present** | 2469 | 3.0 | 1281 | 2.9 | 547 | 3.7 | 485 | 2.5 | 156 | 4.5 |
| **Carotid Procedure** | **Absent** | 80010 | 98.2 | 43488 | 98.5 | 14285 | 97.8 | 18875 | 97.8 | 3362 | 98.2 |
| | **Present** | 1473 | 1.8 | 670 | 1.5 | 320 | 2.2 | 422 | 2.2 | 61 | 1.8 |
| **Percutaneous Intervention** | **Absent** | 78104 | 95.9 | 42504 | 96.3 | 13872 | 95.0 | 18460 | 95.7 | 3268 | 95.5 |
| | **Present** | 3368 | 4.1 | 1638 | 3.7 | 737 | 5.0 | 839 | 4.3 | 154 | 4.5 |
| **APOE2 Carrier** | **Absent** | 133941 | 87.1 | 64812 | 84.7 | 23348 | 87.5 | 40117 | 91.3 | 5664 | 84.8 |
| | **Present** | 19867 | 12.9 | 11686 | 15.3 | 3349 | 12.5 | 3815 | 8.7 | 1017 | 15.2 |
| **APOE4 Carrier** | **Absent** | 95321 | 62.0 | 54181 | 70.8 | 16030 | 60.0 | 20424 | 46.5 | 4686 | 70.1 |
| | **Present** | 58487 | 38.0 | 22317 | 29.2 | 10667 | 40.0 | 23508 | 53.5 | 1995 | 29.9 |
| **CANCER** | **Absent** | 66513 | 81.6 | 35978 | 81.4 | 11775 | 80.5 | 16079 | 83.3 | 2681 | 77.9 |
| | **Present** | 15046 | 18.4 | 8212 | 18.6 | 2846 | 19.5 | 3226 | 16.7 | 762 | 22.1 |
| **Congestive Heart Failure** | **Absent** | 79647 | 97.6 | 43350 | 98.1 | 14196 | 97.1 | 18798 | 97.2 | 3303 | 96.1 |
| | **Present** | 1946 | 2.4 | 845 | 1.9 | 417 | 2.9 | 550 | 2.8 | 134 | 3.9 |
| **Type I Diabetes** | **Absent** | 80722 | 99.5 | 43699 | 99.6 | 14436 | 99.3 | 19182 | 99.5 | 3405 | 99.6 |
| | **Present** | 423 | 0.5 | 195 | 0.4 | 109 | 0.7 | 105 | 0.5 | 14 | 0.4 |
| **Type II Diabetes** | **Absent** | 70745 | 87.2 | 38573 | 87.9 | 12258 | 84.3 | 17013 | 88.2 | 2901 | 84.8 |
| | **Present** | 10400 | 12.8 | 5321 | 12.1 | 2287 | 15.7 | 2274 | 11.8 | 518 | 15.2 |
| **Hearing Aids** | **Absent** | 123764 | 81.6 | 61067 | 81.4 | 22661 | 78.8 | 34391 | 84.0 | 5645 | 80.8 |
| | **Present** | 27912 | 18.4 | 13912 | 18.6 | 6102 | 21.2 | 6557 | 16.0 | 1341 | 19.2 |
| **Hearing Functional** | **Absent** | 17168 | 11.4 | 7064 | 9.5 | 3555 | 12.4 | 5489 | 13.5 | 1060 | 15.3 |
| | **Present** | 133171 | 88.6 | 67217 | 90.5 | 25030 | 87.6 | 35044 | 86.5 | 5880 | 84.7 |
| **Hearing Loss** | **Absent** | 40159 | 26.6 | 18596 | 25.0 | 8711 | 30.3 | 10720 | 26.3 | 2132 | 30.6 |
| | **Present** | 110846 | 73.4 | 55923 | 75.0 | 20023 | 69.7 | 30063 | 73.7 | 4837 | 69.4 |
| **Hispanic** | **No** | 165094 | 93.1 | 80713 | 94.2 | 29190 | 92.1 | 48162 | 92.3 | 7029 | 90.3 |
| | **Yes** | 12253 | 6.9 | 4990 | 5.8 | 2493 | 7.9 | 4015 | 7.7 | 755 | 9.7 |
| **Heart Valve Repair** | **Absent** | 80250 | 98.6 | 43511 | 98.8 | 14314 | 98.0 | 19063 | 98.7 | 3362 | 98.2 |
| | **Present** | 1135 | 1.4 | 533 | 1.2 | 298 | 2.0 | 243 | 1.3 | 61 | 1.8 |
| **Hyper-Cholesteremia** | **Absent** | 35566 | 43.8 | 19607 | 44.6 | 5800 | 39.9 | 8690 | 45.3 | 1469 | 42.8 |
| | **Present** | 45546 | 56.2 | 24337 | 55.4 | 8734 | 60.1 | 10514 | 54.7 | 1961 | 57.2 |
| **Hypertension** | **Absent** | 39769 | 48.7 | 22259 | 50.4 | 6312 | 43.2 | 9792 | 50.6 | 1406 | 40.9 |
| | **Present** | 41846 | 51.3 | 21944 | 49.6 | 8316 | 56.8 | 9558 | 49.4 | 2028 | 59.1 |
| **Hyposomnia/ Insomnia** | **Absent** | 68939 | 85.3 | 36918 | 84.6 | 12112 | 83.1 | 17156 | 89.5 | 2753 | 80.7 |
| | **Present** | 11840 | 14.7 | 6719 | 15.4 | 2455 | 16.9 | 2008 | 10.5 | 658 | 19.3 |
| **Extensive White Matter Hyperintensities** | **Absent** | 32800 | 96.9 | 14384 | 98.5 | 7474 | 96.2 | 9771 | 95.1 | 1171 | 97.3 |
| | **Present** | 1042 | 3.1 | 214 | 1.5 | 293 | 3.8 | 502 | 4.9 | 33 | 2.7 |
| **Lacunar Infarcts** | **Absent** | 30166 | 90.0 | 13055 | 92.2 | 6816 | 87.7 | 9253 | 89.2 | 1042 | 86.3 |
| | **Present** | 3351 | 10.0 | 1111 | 7.8 | 957 | 12.3 | 1118 | 10.8 | 165 | 13.7 |
| **Large Vessel Infarcts** | **Absent** | 32584 | 97.8 | 13912 | 98.9 | 7516 | 97.1 | 9988 | 97.0 | 1168 | 96.6 |
| | **Present** | 723 | 2.2 | 152 | 1.1 | 224 | 2.9 | 306 | 3.0 | 41 | 3.4 |
| **Macro-hemorrhages** | **Absent** | 32227 | 99.3 | 13669 | 99.5 | 7443 | 99.1 | 9931 | 99.1 | 1184 | 99.4 |
| | **Present** | 243 | 0.7 | 74 | 0.5 | 71 | 0.9 | 91 | 0.9 | 7 | 0.6 |
| **Micro-hemorrhages** | **Absent** | 23911 | 91.2 | 10248 | 93.1 | 5614 | 89.9 | 7188 | 89.9 | 861 | 88.9 |
| | **Present** | 2308 | 8.8 | 762 | 6.9 | 629 | 10.1 | 809 | 10.1 | 108 | 11.1 |
| **Moderate White Matter Hyperintensities** | **Absent** | 29222 | 85.8 | 13175 | 89.7 | 6475 | 82.6 | 8536 | 82.7 | 1036 | 85.8 |
| | **Present** | 4833 | 14.2 | 1506 | 10.3 | 1368 | 17.4 | 1788 | 17.3 | 171 | 14.2 |
| **Marital Status** | **divorced** | 20596 | 11.6 | 12166 | 14.2 | 3629 | 11.5 | 3697 | 7.1 | 1104 | 14.2 |
| | **domestic partner** | 2697 | 1.5 | 1483 | 1.7 | 498 | 1.6 | 618 | 1.2 | 98 | 1.3 |
| | **married** | 107123 | 60.5 | 47942 | 56.0 | 19094 | 60.4 | 35670 | 68.4 | 4417 | 56.9 |
| | **never married** | 8401 | 4.7 | 4966 | 5.8 | 1571 | 5.0 | 1368 | 2.6 | 496 | 6.4 |
| | **separated** | 1447 | 0.8 | 643 | 0.8 | 345 | 1.1 | 343 | 0.7 | 116 | 1.5 |
| | **widowed** | 36908 | 20.8 | 18411 | 21.5 | 6490 | 20.5 | 10470 | 20.1 | 1537 | 19.8 |
| **Myocardial Infarction** | **Absent** | 78765 | 96.5 | 42966 | 97.1 | 13957 | 95.4 | 18553 | 95.8 | 3289 | 95.6 |
| | **Present** | 2897 | 3.5 | 1270 | 2.9 | 672 | 4.6 | 805 | 4.2 | 150 | 4.4 |
| **Anxiolytics** | **Absent** | 151429 | 86.3 | 73679 | 87.3 | 26995 | 85.7 | 44251 | 85.6 | 6504 | 83.7 |
| | **Present** | 23939 | 13.7 | 10744 | 12.7 | 4489 | 14.3 | 7435 | 14.4 | 1271 | 16.3 |
| **Anticoagulants** | **Absent** | 100979 | 57.6 | 49340 | 58.4 | 17243 | 54.8 | 30135 | 58.3 | 4261 | 54.8 |
| | **Present** | 74389 | 42.4 | 35083 | 41.6 | 14241 | 45.2 | 21551 | 41.7 | 3514 | 45.2 |
| **Antidepressants** | **Absent** | 124191 | 70.8 | 67909 | 80.4 | 22284 | 70.8 | 28433 | 55.0 | 5565 | 71.6 |
| | **Present** | 51177 | 29.2 | 16514 | 19.6 | 9200 | 29.2 | 23253 | 45.0 | 2210 | 28.4 |
| **Alzheimer's Disease medications** | **Absent** | 132307 | 75.4 | 83316 | 98.7 | 24301 | 77.2 | 17570 | 34.0 | 7120 | 91.6 |
| | **Present** | 43061 | 24.6 | 1107 | 1.3 | 7183 | 22.8 | 34116 | 66.0 | 655 | 8.4 |
| **Anti-hypertensives** | **Absent** | 74215 | 42.3 | 36196 | 42.9 | 11619 | 36.9 | 23480 | 45.4 | 2920 | 37.6 |
| | **Present** | 101153 | 57.7 | 48227 | 57.1 | 19865 | 63.1 | 28206 | 54.6 | 4855 | 62.4 |
| **Familial Alzheimer's Disease Mutations** | **Absent** | 39674 | 99.3 | 20199 | 99.3 | 7661 | 99.3 | 9831 | 99.2 | 1983 | 99.5 |
| | **APP** | 231 | 0.6 | 133 | 0.7 | 43 | 0.6 | 46 | 0.5 | 9 | 0.5 |
| | **PSEN1** | 54 | 0.1 | 15 | 0.1 | 4 | 0.1 | 35 | 0.4 | NA | NA |
| | **PSEN2** | 11 | 0.0 | 3 | 0.0 | 5 | 0.1 | 3 | 0.0 | NA | NA |
| **APOE** | **2/2** | 631 | 0.4 | 378 | 0.5 | 115 | 0.4 | 86 | 0.2 | 52 | 0.8 |
| | **2/3** | 15255 | 9.9 | 9389 | 12.3 | 2484 | 9.3 | 2588 | 5.9 | 794 | 11.9 |
| | **3/3** | 79435 | 51.6 | 44414 | 58.1 | 13431 | 50.3 | 17750 | 40.4 | 3840 | 57.5 |
| | **3/4** | 45605 | 29.7 | 18538 | 24.2 | 8214 | 30.8 | 17182 | 39.1 | 1671 | 25.0 |
| | **2/4** | 3981 | 2.6 | 1919 | 2.5 | 750 | 2.8 | 1141 | 2.6 | 171 | 2.6 |
| | **4/4** | 8901 | 5.8 | 1860 | 2.4 | 1703 | 6.4 | 5185 | 11.8 | 153 | 2.3 |
| **Antipsychotics** | **Absent** | 167994 | 95.8 | 83792 | 99.3 | 30810 | 97.9 | 45816 | 88.6 | 7576 | 97.4 |
| | **Present** | 7374 | 4.2 | 631 | 0.7 | 674 | 2.1 | 5870 | 11.4 | 199 | 2.6 |
| **Diabetes Medications** | **Absent** | 156438 | 89.2 | 75705 | 89.7 | 27472 | 87.3 | 46462 | 89.9 | 6799 | 87.4 |
| | **Present** | 18930 | 10.8 | 8718 | 10.3 | 4012 | 12.7 | 5224 | 10.1 | 976 | 12.6 |
| **Estrogen Hormone Therapy** | **Absent** | 169512 | 96.7 | 80434 | 95.3 | 30670 | 97.4 | 50871 | 98.4 | 7537 | 96.9 |
| | **Present** | 5856 | 3.3 | 3989 | 4.7 | 814 | 2.6 | 815 | 1.6 | 238 | 3.1 |
| **Estrogen + Progesterone Therapy** | **Absent** | 174848 | 99.7 | 84072 | 99.6 | 31418 | 99.8 | 51610 | 99.9 | 7748 | 99.7 |
| | **Present** | 520 | 0.3 | 351 | 0.4 | 66 | 0.2 | 76 | 0.1 | 27 | 0.3 |

| | | | | | | | | | | | |
|---|---|---|---|---|---|---|---|---|---|---|---|
| **Family History of Cognitive Impairment** | **Absent** | 62361 | 38.5 | 30101 | 38.1 | 11472 | 39.9 | 17864 | 38.1 | 2924 | 41.0 |
| | **Present** | 99423 | 61.5 | 48910 | 61.9 | 17265 | 60.1 | 29032 | 61.9 | 4216 | 59.0 |
| **Lipid Lowering Medications** | **Absent** | 93037 | 53.1 | 45249 | 53.6 | 15338 | 48.7 | 28510 | 55.2 | 3940 | 50.7 |
| | **Present** | 82331 | 46.9 | 39174 | 46.4 | 16146 | 51.3 | 23176 | 44.8 | 3835 | 49.3 |
| **NIH Race Categories** | **American Indian/Alaska Native** | 709 | 0.4 | 332 | 0.4 | 133 | 0.4 | 210 | 0.4 | 34 | 0.4 |
| | **Asian** | 4377 | 2.5 | 2188 | 2.6 | 947 | 3.0 | 1065 | 2.1 | 177 | 2.3 |
| | **Biracial** | 4882 | 2.8 | 2312 | 2.7 | 1014 | 3.2 | 1323 | 2.6 | 233 | 3.1 |
| | **Black** | 21775 | 12.4 | 11692 | 13.7 | 4323 | 13.8 | 4519 | 8.8 | 1241 | 16.3 |
| | **Caucasian** | 143860 | 81.9 | 68802 | 80.6 | 24987 | 79.5 | 44157 | 86.1 | 5914 | 77.8 |
| | **Native Hawaiin/Pacific Islander** | 95 | 0.1 | 48 | 0.1 | 12 | 0.0 | 29 | 0.1 | 6 | 0.1 |
| **Non-steroidal Anti-Inflammatory Drugs** | **Absent** | 99969 | 57.0 | 46873 | 55.5 | 17606 | 55.9 | 31210 | 60.4 | 4280 | 55.0 |
| | **Present** | 75399 | 43.0 | 37550 | 44.5 | 13878 | 44.1 | 20476 | 39.6 | 3495 | 45.0 |
| **Parkinson's Disease Medications** | **Absent** | 167035 | 95.2 | 81333 | 96.3 | 29767 | 94.5 | 48571 | 94.0 | 7364 | 94.7 |
| | **Present** | 8333 | 4.8 | 3090 | 3.7 | 1717 | 5.5 | 3115 | 6.0 | 411 | 5.3 |
| **Osteoarthritis** | **Absent** | 33991 | 43.4 | 16380 | 38.5 | 6005 | 42.7 | 10341 | 56.2 | 1265 | 38.5 |
| | **Present** | 44279 | 56.6 | 26127 | 61.5 | 8070 | 57.3 | 8064 | 43.8 | 2018 | 61.5 |
| **Pacemaker Implantation** | **Absent** | 79038 | 97.1 | 43083 | 97.8 | 14066 | 96.2 | 18579 | 96.2 | 3310 | 96.7 |
| | **Present** | 2379 | 2.9 | 975 | 2.2 | 550 | 3.8 | 740 | 3.8 | 114 | 3.3 |
| **REM Behavior Disorder** | **Absent** | 76512 | 95.9 | 42267 | 98.3 | 13579 | 94.2 | 17452 | 92.0 | 3214 | 95.3 |
| | **Present** | 3257 | 4.1 | 750 | 1.7 | 838 | 5.8 | 1512 | 8.0 | 157 | 4.7 |
| **Rheumatoid Arthritis** | **Absent** | 76962 | 98.3 | 41808 | 98.4 | 13826 | 98.2 | 18119 | 98.4 | 3209 | 97.7 |
| | **Present** | 1308 | 1.7 | 699 | 1.6 | 249 | 1.8 | 286 | 1.6 | 74 | 2.3 |
| **Seizure History** | **Absent** | 108630 | 97.2 | 47935 | 98.2 | 20697 | 97.2 | 35161 | 96.0 | 4837 | 96.3 |
| | **Present** | 3143 | 2.8 | 886 | 1.8 | 588 | 2.8 | 1484 | 4.0 | 185 | 3.7 |
| **Sleep Apnea** | **Absent** | 63884 | 80.5 | 34983 | 81.6 | 10949 | 76.7 | 15337 | 81.1 | 2615 | 77.7 |
| | **Present** | 15520 | 19.5 | 7868 | 18.4 | 3322 | 23.3 | 3579 | 18.9 | 751 | 22.3 |
| **Thyroid Disease** | **Absent** | 64550 | 79.8 | 34599 | 79.2 | 11702 | 80.7 | 15530 | 80.9 | 2719 | 79.4 |
| | **Present** | 16294 | 20.2 | 9108 | 20.8 | 2805 | 19.3 | 3676 | 19.1 | 705 | 20.6 |
| **Tobacco Use** | **Absent** | 108067 | 96.6 | 47287 | 96.8 | 20525 | 96.4 | 35480 | 96.8 | 4775 | 95.1 |
| | **Present** | 3748 | 3.4 | 1545 | 3.2 | 775 | 3.6 | 1183 | 3.2 | 245 | 4.9 |
| **Vitamin B12 Deficiency** | **Absent** | 73710 | 92.9 | 40326 | 94.3 | 13070 | 91.8 | 17188 | 90.7 | 3126 | 92.5 |
| | **Present** | 5620 | 7.1 | 2437 | 5.7 | 1170 | 8.2 | 1758 | 9.3 | 255 | 7.5 |
| **Gender** | **Female** | 103371 | 58.1 | 56106 | 65.2 | 16300 | 51.3 | 26536 | 50.7 | 4429 | 56.8 |
| | **Male** | 74589 | 41.9 | 29887 | 34.8 | 15500 | 48.7 | 25832 | 49.3 | 3370 | 43.2 |

**Supplementary Table 3: Demographics and Clinical Variables by Cognitive Status**. Demographic and clinical variables were analyzed for each cognitive status, including Impaired-not-MCI (not included in the rest of the analyses). For continuous variables, the mean and standard deviation are reported. For categorical variables, the overall count and frequency per category are reported.
